# Maternal postnatal depression and offspring emotional and behavioural development at age 7 years in a UK-birth cohort: the role of paternal involvement

**DOI:** 10.1101/2021.10.12.21264846

**Authors:** Iryna Culpin, Gemma Hammerton, Alan Stein, Marc H Bornstein, Henning Tiemeier, Tim Cadman, Eivor Fredriksen, Jonathan Evans, Tina Miller, Esther Dermott, Jon Heron, Hannah M Sallis, Rebecca M Pearson

## Abstract

**Background:** There is considerable variability in emotional and behavioural outcomes of children whose mothers experience depression. Few longitudinal studies have examined potential contributions of dimensions of paternal involvement in the association between maternal postnatal depression (PND) and offspring development.

**Methods:** We examined pathways from maternal PND at 8 weeks postnatally (assessed using the Edinburgh Postnatal Depression Scale) to offspring emotional and behavioural development at 7 years (assessed using the Strengths and Difficulties Questionnaire) through behavioural, affective and cognitive child-focused and mother-influenced dimensions of paternal involvement in 3,434 members of the UK-based birth cohort, the Avon Longitudinal Study of Parents and Children. Analyses were adjusted for a range of baseline confounders and paternal postnatal depression (PND) as an intermediate confounder.

**Results:** Maternal PND was associated with higher levels of some aspects of child-focused and mother-influenced paternal involvement in models accounting for paternal PND, however these pathways were not associated with offspring emotional and behavioural development at age 7 years. There was strong evidence of direct effect from maternal PND to offspring development, but no evidence of mediation through the combination of all indirect pathways through child-focused and mother-influenced paternal involvement. However, higher levels of father-child conflict were associated with increased risk of offspring emotional and behavioural difficulties, and this pathway mediated a proportion of the maternal PND to offspring risk. Additionally, maternal PND was associated with paternal PND, which, in turn, was associated with lower levels of child-focused and mother-influenced paternal involvement.

**Conclusions:** The positive associations between maternal PND and some aspects of paternal involvement suggest that non-depressed fathers may engage in ‘compensatory’ parenting strategies in response to maternal PND, which although important may not be sufficient in reducing the adverse impact of maternal PND on offspring emotional and behavioural development. Conflictual father-child relationships emerged as a risk factor for adverse offspring development and as an explanatory mechanism in the association between maternal PND and offspring development. These results suggest that interventions that reduce father-child conflict may reduce the risk of emotional and behavioural difficulties in offspring of depressed mothers.

## Introduction

The associations between maternal postnatal depression (PND) and adverse offspring emotional and behavioural development across the lifespan are well-established.^1^ However, outcomes of children whose mothers experience PND are not consistently poor.^1^ Thus, it is important to understand what processes underly both resilience and adverse offspring outcomes.^2^ Insights into such mechanisms may improve our understanding of intergenerational transmission of mental health risks in families and contribute to the development of targeted interventions to reduce adverse impact of maternal PND on offspring development. Ample evidence suggests that quality of maternal parenting is one important mechanism of transmission,^1,3^ with maternal PND disrupting sensitivity and involvement,^4^ which, in turn, are associated with offspring emotional and behavioural problems.^5^ However, few longitudinal studies have examined the role that paternal involvement plays in the association between maternal PND and offspring development,^6^ with most existing studies addressing fathers’ role in the context of ‘normative’ family functioning.^7-10^

Both ‘compensatory/buffering’ and ‘spillover’ hypotheses have been articulated to describe the role of fathers in the context of maternal PND.^6^ In line with family-level resilience perspective,^11-12^ fathers may try to compensate for maternal PND by becoming more involved, ‘buffering’ the adverse effects of maternal PND on the child.^2^ However, maternal PND may have a ‘spillover’ effect by negatively affecting paternal involvement and increasing the risk of adverse offspring outcomes.^6^ Evidence for ‘compensatory/buffering’ effects of paternal involvement on offspring outcomes is inconsistent. Some studies report an increase in positive, warm and sensitive father-child interactions^13-15^ in families with depressed compared to non-depressed mothers,^2,15^ but other studies report no evidence that paternal involvement is protective (i.e., moderates the association between maternal PND and offspring development).^16,17^ The ‘spillover’ hypothesis has also been substantiated with some evidence linking maternal PND to lower levels of paternal involvement with their infants.^18^ However, there remains paucity of evidence regarding the impact of maternal PND on the nature and quality of paternal involvement, and, to the best of our knowledge, no large population-based studies to-date have examined a possible explanatory (i.e., mediating) role of paternal involvement in the context of maternal PND. This is an important omission given growing evidence emphasising the importance of paternal involvement for numerous aspects of offspring development^19,20^ and a call for an integrated (i.e., inclusive of father) scientific and clinical approach to family functioning affected by maternal PND.^11,12^

Paternal involvement is a multidimensional construct, with conceptual definitions long departing from traditional breadwinning roles to ‘intimate’ fatherhood that prioritises emotional closeness and the quality of the parent-child relationship.^21^ Paternal involvement is also no longer solely defined as time spent together,^20^ with more nuanced facets, including direct nurturant caregiving and disciplining as well as emotional and practical support of the mother, being linked to improved emotional and behavioural outcomes in offspring.^9^ New conceptualisations of paternal involvement broadly include child-focused behavioural (e.g., direct involvement in caregiving), affective (e.g., quality of father-child relationship, such as enjoyment, warmth and conflict), and cognitive (e.g., parenting confidence and beliefs regarding caregiving) dimensions^22-24^ as well as aspects of involvement that are shaped by maternal influences and mother-father relationship (e.g., ‘gatekeeping’, a complex phenomenon defined as maternal beliefs and practices that discourage or facilitate paternal involvement).^25-27^ Supporting the mother through sharing household tasks and responsibilities is another important dimension of paternal involvement, with significant consequences for the child.^22^ Similar to mothers, fathers are also under pressure to balance caregiving with demands of paid work, childcare and family life,^28,29^ often accompanied by mothers’ perceptions that fathers are not ‘pulling their fair share’ with this benchmarking influencing paternal inolvement.^26^ The recognition that child-focused and mother-influenced paternal involvement are important for offspring development is another marked shift in fatherhood scholarship,^9^ explicitly acknowledging that father-child relationship is embedded in a network of relationships among fathers, mothers and children.^30,31^ However, few studies have examined paternal involvement as a multidimensional construct that captures cognitive, affective and behavioural aspects as well as maternal influences.^31,32^ Failures to do so preclude establishing the individual relative contributions of various aspects of paternal involvement to offspring outcomes.^9^ We address this limitation by examining several dimensions of child-focused and mother-influenced paternal involvement in relation to offspring emotional and behavioural outcomes in the context of maternal PND.

Parenting is transactional and dynamic.^33,34^ Existing research acknowledges reciprocal influences and the interplay between paternal involvement and the child, with offspring response to parenting shaped by temperamental and biological dispositions, which may evoke variations in parenting behaviours.^35,36^ In addition, paternal PND and its impact on offspring development in the context of maternal PND is often overlooked. Maternal depression in pregnancy and during the postnatal period is associated with increased risk of paternal PND,^37,38^ which in turn influences paternal parenting,^39^ and offspring emotional and behavioural development,^40^ thus potentially influencing the association between paternal involvement (mediator) and offspring development (outcome). Figure 1 presents conceptual pathways between maternal and paternal PND, paternal involvement and offspring development. Paternal PND co-occuring with maternal PND may increase offspring risk of adverse development,^6^ while absence of paternal PND may be a protective factor reducing adverse effects of maternal PND on paternal involvement and offspring outcomes. Mezulis et al.^2^ found that paternal PND exacerbated adverse effects of maternal PND on offspring behavioural problems, but this moderating effect was limited to depressed fathers spending considerable amounts of time caring for their children. To the best of our knowledge, no longitudinal study has examined the role of paternal PND in the association between paternal involvement and offspring development.

**Figure 1.**
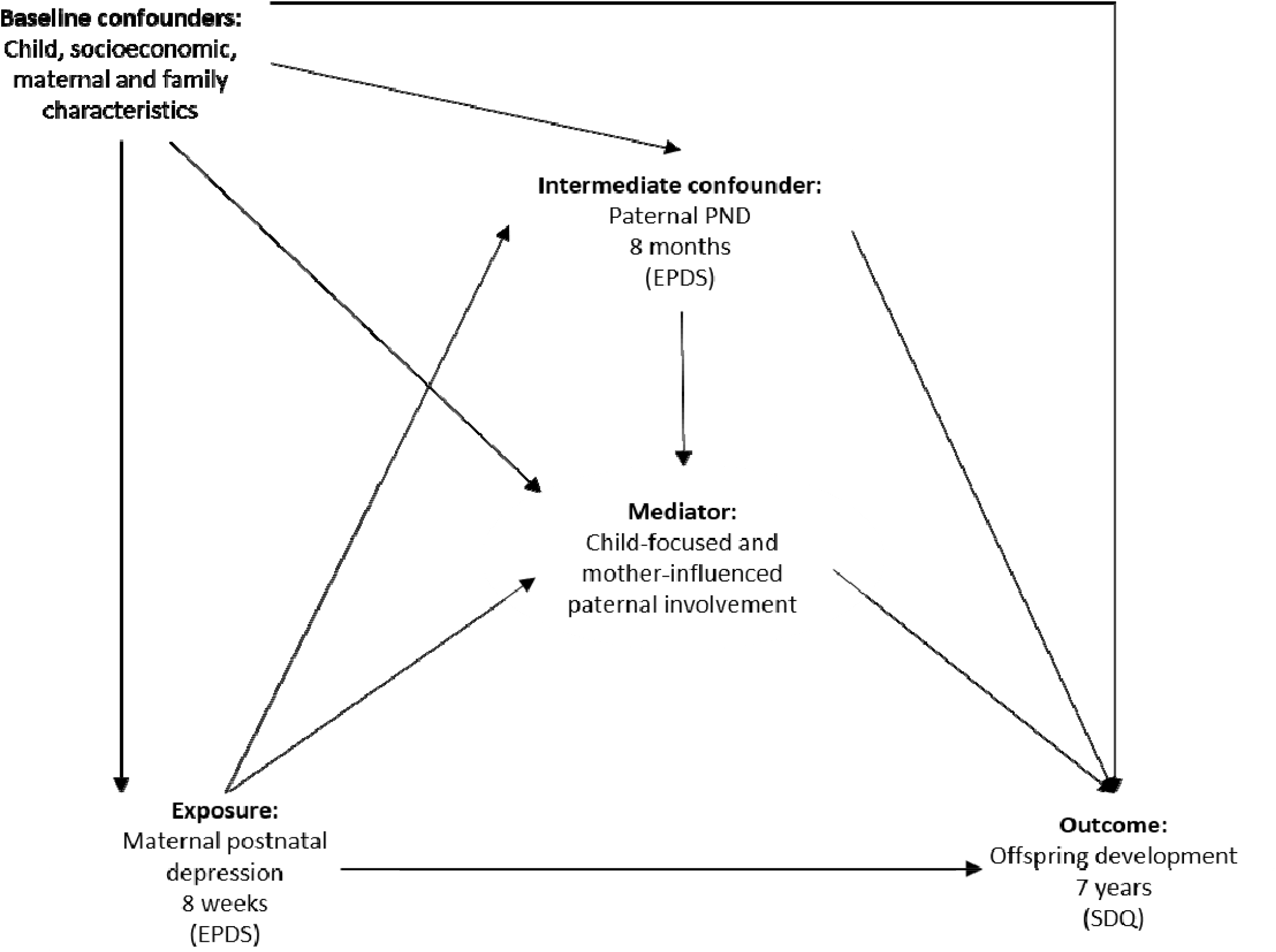
Conceptual diagram for exposure (maternal PND), mediator (child-focused and mother-influenced dimensions of paternal involvement), outcome (offspring development), baseline confounders (child, socioeconomic, maternal and family characteristics) and intermediate confounder * (paternal PND) ^*^ Intermediate confounder: variable induced by the exposure, affecting both the mediator and the outcome, thus confounding mediator-outcome association

In the present study, we took advantage of rich self-reported longitudinal data on paternal parenting collected as part of a large UK-based birth cohort study, the Avon Longitudinal Study of Parents and Children (ALSPAC) to (1) model child-focused and mother-influenced dimensions of paternal engagement and (2) estimate the extent to which the association between maternal PND and offspring emotional and behavioural difficulties at 7 years is explained (mediated) by child-focused and mother-influenced dimensions of paternal involvement during the first four years of the child’s life. Quantifying this association has important implications for the design and development of preventative and intervention programmes, given the modifiable nature of parenting.^41^ The unique nature of the rich longitudinal ALSPAC data enabled us also to account for a range of confounding factors, including child polygenic score for neuroticism (PGS) as a proxy for genetic confounding that may indicate possible child evocative effects, and paternal PND as a possible intermediate confounder.

## Methods

### Study cohort

The sample comprised participants from the ALSPAC cohort. During Phase I enrolment, 14,541 pregnant mothers residing in the former Avon Health Authority in the south-west of England with expected dates of delivery between 1 April 1991 and 31 December 1992 were recruited. The total sample size is 15,454 pregnancies, of which 14,901 were alive at 1 year of age. Our sample comprised 9,766 fathers with at least one parenting item. Ethical approval and informed consent for the data collection were obtained from the ALSPAC Ethics and Law Committee and the Local Research Ethics Committees (http://www.bristol.ac.uk/alspac/researchers/research-ethics/). Information about ALSPAC is available at www.bristol.ac.uk/alspac/, including a fully searchable data dictionary (http://www.bris.ac.uk/alspac/researchers/our-data/). Further details on the cohort profile, representativeness and phases of recruitment are described in three cohort-profile papers.^42-44^

### Measures

#### Exposure: maternal postnatal depression

Symptoms of maternal PND were measured at 8 weeks postnatally using the Edinburgh Postnatal Depression Scale (EPDS),^45^ a 10-item self-reported depression questionnaire validated for use during the perinatal period. The earliest postnatal time point was chosen to align with the earliest assessment of paternal involvement items at 8 weeks. To make full use of variation in symptoms individual depression items were summed to derive a continuous score (range 0-30).^46^ The continuous score was used in all analyses, except descriptive characteristics of the study sample by exposure status (threshold ≥13; 26.6%; Table S1, Supplementary).

#### Outcome: offspring emotional and behavioural development

Offspring emotional and behavioural development was assessed using the Strengths and Difficulties Questionnaire^47^ completed by the mothers of the study children at age 81 months (hereafter referred to as 7 years). The scale consists of 25 questions with five subscales (prosocial [i.e., engagement in positive social interactions], emotional symptoms, conduct problems, hyperactivity and peer problems), which have been extensively validated, demonstrating high consistency, reliability and diagnostic predictability.^48^ Consistent with previous research, the first 4 subscales were combined to derive a total difficulties score used in all analyses.^49^

#### Mediators: child-focused and mother-influenced dimensions of paternal involvement

Full details pertaining to item selection and theoretical development of factors are presented in Methods S1 (Supplementary). In summary, potential parenting items (>150) were extracted from paternal self-reported questionnaires administered on 5 occasions from birth to age 3 years 11 months. Conceptual factor underpinnings were drawn from extensive empirical and sociological literature on paternal engagement in infancy.^19,24,26,31,50^ In line with revised conceptualisation of paternal involvement,^26,31,32^ individual items fell into two theoretically distinct sources of paternal involvement: (1) *child-focused* paternal involvement capturing behavioural (e.g., direct involvement in caregiving), affective (e.g., quality of father-child relationship, such as enjoyment, warmth and conflict, worries about the child) and cognitive (e.g., parenting confidence and beliefs regarding caregiving) dimensions directed at the child; and (2) *mother-influenced* paternal involvement with the child through the lens of maternal expectations (e.g., maternal ‘gatekeeping’, managing employment and parenthood), mother-father relationship (e.g., paternal beliefs regarding mother-father relationship and its impact on parenting) and indirect material care through support of the mother (e.g., paternal help with household tasks and responsibilities). The hypothesised latent factor (CFA) models capturing child-focused (6 dimensions) and mother-influenced (4 dimensions) paternal involvement are represented in Figure 2a-b, with derived factors, items, standardised factor loadings and fit indices for the measurement models presented in Tables 1-2. Two separate measurement models including CFA-based factors (Figure 1a-b) and structural mediation models including pathways between constructs (Figure 3a-b) were fitted to capture potentially differential roles of child-focused and mother-influenced paternal involvement in the association between maternal PND and offspring development.

**Table 1.** Derived factors, items, standardised factor loadings and fit indices for the measurement model capturing child-focused dimensions of paternal involvement

| Item | Age at assessment/Factor | Item description | Factor loading | S.E. |
| --- | --- | --- | --- | --- |
| **I** | **Paternal parenting confidence** |  |  |  |
| 1. | 8 weeks | Partner feels confident with child | 0.521 | 0.016 |
| 2. | 8 weeks | Partner happy with the way he brings up child | 0.657 | 0.015 |
| 3. | 8 weeks | Partner regrets lack of experience with child | 0.363 | 0.022 |
| 4. | 8 weeks | Partner feels well-prepared for birth and childcare | 0.403 | 0.015 |
| 5. | 8 weeks | Having a baby is as expected | 0.356 | 0.015 |
| 6. | 8 months | Partner feels confident with child | 0.711 | 0.021 |
| 7. | 8 months | Partner unsure whether doing the right thing | 0.436 | 0.017 |
| 8. | 1 year 9 months | Partner afraid to be alone with toddler | 0.478 | 0.055 |
| 9. | 1 year 9 months | Partner sure doing the right thing for child | 0.574 | 0.016 |
| 10. | 2 years 9 months | Partner feels confident with child | 0.694 | 0.021 |
| 11. | 2 years 9 months | Partner feels constantly unsure if doing right thing for child | 0.434 | 0.019 |
| **II** | **Paternal conflictual relationship with child** |  |  |  |
| 1. | 8 weeks | Partner so stressed at home it is a bad influence on child | 0.557 | 0.017 |
| 2. | 8 weeks | Partner regrets having child | 0.688 | 0.022 |
| 3. | 8 months | Partner dislikes mess surrounding child | 0.532 | 0.014 |
| 4. | 8 months | Partner finds child crying unbearable | 0.564 | 0.014 |
| 5. | 8 months | Partner feels they have no time to themselves | 0.590 | 0.013 |
| 6. | 1 year 9 months | Child whining can make parent want to hit | 0.477 | 0.015 |
| 7. | 1 year 9 months | Having young child is absolutely exhausting | 0.451 | 0.014 |
| 8. | 1 year 9 months | Smacking is the best way to discipline child | 0.168 | 0.017 |
| 9. | 1 year 9 months | Parent can feel exasperated calming child | 0.366 | 0.015 |
| 10. | 1 year 9 months | Child never gets on partner's nerves | 0.306 | 0.015 |
| 11. | 1 year 9 months | Partner cannot bear it when child cries | 0.364 | 0.016 |
| 12. | 1 year 9 months | Partner feels desperate when child complains | 0.548 | 0.015 |
| 13. | 1 year 9 months | Child demands bring intense anger | 0.555 | 0.017 |
| 14. | 2 years 9 months | Partner dislikes mess surrounding child | 0.550 | 0.015 |
| 15. | 2 years 9 months | Partner cannot bear it when child cries | 0.434 | 0.016 |
| 16. | 2 years 9 months | Partner feels they have no time to themselves | 0.556 | 0.015 |
| 17. | 2 years 9 months | Partner often gets very irritated with child | 0.454 | 0.020 |
| 18. | 2 years 9 months | Partner has frequent battles of will with child | 0.290 | 0.021 |
| 19. | 2 years 9 months | Child gets on partner's nerves | 0.438 | 0.025 |
| **III** | **Paternal enjoyment and warmth** |  |  |  |
| 1. | 8 weeks | Partner is making a strong bond with child | 0.515 | 0.013 |
| 2. | 8 weeks | Partner feels guilty at not enjoying child | 0.651 | 0.015 |
| 3. | 8 weeks | Child made partner more fulfilled | 0.486 | 0.013 |
| 4. | 8 months | Partner enjoys child | 0.657 | 0.014 |
| 5. | 8 months | Partner preferred not to have had child | 0.559 | 0.018 |
| 6. | 8 months | Pleasure watching child develop | 0.693 | 0.016 |
| 7. | 8 months | Partner feels should enjoy child, but they are not | 0.704 | 0.015 |
| 8. | 8 months | Child made partner more fulfilled | 0.550 | 0.012 |
| 9. | 8 months | Partner feels children are fun | 0.646 | 0.012 |
| 10. | 8 months | Partner enjoys seeing child after work | 0.639 | 0.016 |
| 11. | 8 months | Partners feels having child made more fulfilled | 0.541 | 0.016 |
| 12. | 8 months | Talking to child is important | 0.352 | 0.036 |
| 13. | 8 months | Cuddling child is very important | 0.375 | 0.037 |
| 14. | 1 year 9 months | Children are fun | 0.568 | 0.014 |
| 15. | 1 year 9 months | Partner really loves child | 0.708 | 0.020 |
| 16. | 1 year 9 months | Partner glad they had child | 0.670 | 0.016 |
| 17. | 1 year 9 months | Partner feels great pleasure watching child grow | 0.673 | 0.018 |
| 18. | 1 year 9 months | Child gives great joy | 0.705 | 0.016 |
| 19. | 2 years 9 months | Partner really enjoys child | 0.659 | 0.015 |
| 20. | 2 years 9 months | Partner feels great pleasure watching child develop | 0.730 | 0.017 |
| 21. | 2 years 9 months | Partner feels should enjoy child, but they are not | 0.604 | 0.017 |
| 22. | 2 years 9 months | Partners feels having child made more fulfilled | 0.597 | 0.014 |
| 23. | 2 years 9 months | Partner enjoys seeing child after work | 0.607 | 0.017 |
| 24. | 3 years 11 months | Child makes partner feel very happy | 0.286 | 0.062 |
| 25. | 3 years 11 months | Partner feels very close to child | 0.422 | 0.045 |
| 26. | 3 years 11 months | Child is very affectionate to partner | 0.384 | 0.037 |
| 27. | 3 years 11 months | Cuddling is best way to calm child | 0.643 | 0.015 |
| **IV Paternal involvement in childcare** |  |  |  |  |
| 1. | 8 weeks | Frequency partner changes nappy per week | 0.644 | 0.014 |
| 2. | 8 weeks | Frequency partner bathes child per week | 0.387 | 0.014 |
| 3. | 8 weeks | Frequency partner plays with child per week | 0.751 | 0.022 |
| 4. | 8 weeks | Frequency partner walks child outside per week | 0.220 | 0.014 |
| 5. | 8 weeks | Frequency partner puts child to bed per week | 0.593 | 0.013 |
| 6. | 8 weeks | Frequency partner feeds child or helps at night per week | 0.515 | 0.014 |
| 7. | 8 weeks | Frequency partner feeds child per week | 0.642 | 0.017 |
| 8. | 8 months | Frequency partner gives help changing nappies | 0.449 | 0.020 |
| **V Paternal worries about child** |  |  |  |  |
| 1. | 1 year 9 months | Partner worries if child is eating enough | 0.733 | 0.018 |
| 2. | 1 year 9 months | Get child to eat right food makes partner anxious | 0.935 | 0.021 |
| 3. | 1 year 9 months | Partner anxious if someone else looks after child | 0.372 | 0.017 |
| 4. | 2 years 9 months | Partner worries about study child when at work | 0.340 | 0.024 |
| **VI Paternal beliefs regarding caregiving** |  |  |  |  |
| 1. | 8 months | Babies should be picked up when cry | 0.482 | 0.030 |
| 2. | 8 months | Babies should be fed when hungry | 0.552 | 0.033 |
| 3. | 8 months | Parents need to adapt lives to babies needs | 0.320 | 0.026 |
| 4. | 8 months | Baby should fit into parents' routine | 0.143 | 0.026 |
| 5. | 8 months | Babies should be stimulated to develop well | 0.311 | 0.029 |
| 6. | 1 year 9 months | Toddlers should be able to eat when they ask | 0.207 | 0.042 |
| **Latent factor model fit indices** |  |  |  |  |
| Free parameters |  |  | 299 |  |
| Root Mean Square Error of Approximation (RMSEA) |  |  | 0.022 (95%CI |  |
|  |  |  | 0.021 to 0.022) |  |
| Comparative Fit Index (CFI) |  |  | 0.904 |  |
| Tucker-Lewis Index (TLI) |  |  | 0.899 |  |

**Table 2.** Derived factors, items, standardised factor loadings and fit indices for the measurement model capturing mother-influenced dimensions of paternal involvement

| Item | Age at assessment/Factor | Item description | Factor loading | S.E. |
| --- | --- | --- | --- | --- |
| **I** | **Paternal help with household tasks** |  |  |  |
| 1. | 8 weeks | Partner helped with shopping since birth | 0.518 | 0.011 |
| 2. | 8 weeks | Partner helped with cleaning home since birth | 0.591 | 0.011 |
| 3. | 8 weeks | Partner helped with meal preparation since birth | 0.599 | 0.013 |
| 4. | 8 weeks | Partner helped with washing up since birth | 0.687 | 0.011 |
| 5. | 8 weeks | Partner helped with housework since birth | 0.642 | 0.009 |
| 6. | 8 weeks | Partner helped with cooking meals since birth | 0.613 | 0.010 |
| 7. | 8 weeks | Partner helped with clothes wash since birth | 0.491 | 0.011 |
| 8. | 8 months | Partner gave help with shopping | 0.427 | 0.012 |
| 9. | 8 months | Partner gave help with cleaning home | 0.533 | 0.011 |
| 10. | 8 months | Partner gave help preparing meals | 0.688 | 0.011 |
| 11. | 8 months | Partner gave help washing up | 0.498 | 0.012 |
| 12. | 8 months | Partner gave help washing clothes | 0.414 | 0.012 |
| **II** | **Paternal perception of maternal 'gatekeeping'** |  |  |  |
| 1. | 8 weeks | Pressure on partner to be present at birth | 0.240 | 0.023 |
| 2. | 8 weeks | Mother excludes partner from childcare | 0.660 | 0.020 |
| 3. | 8 weeks | Partner feels mother does not trust him with childcare | 0.696 | 0.016 |
| 4. | 8 weeks | Partner happy with the way mother brings up child | 0.503 | 0.023 |
| 5. | 8 weeks | Home is woman's place, no part for me | 0.649 | 0.024 |
| 6. | 8 weeks | Partner always getting under mother's feet | 0.573 | 0.017 |
| 7. | 8 weeks | Mother dislikes partner being involved with childcare | 0.718 | 0.027 |
| **III** | **Paternal beliefs regarding mother-father relationship and parenting** |  |  |  |
| 1. | 8 weeks | Parenthood has made partner and mother closer | 0.363 | 0.017 |
| 2. | 8 weeks | Mother no longer gives partner attention | 0.709 | 0.018 |
| 3. | 8 weeks | Partner feels hurt by attention mother gives child | 0.758 | 0.024 |
| **IV** | **Paternal beliefs regarding employment and parenthood** |  |  |  |
| 1. | 8 weeks | Mother expects partner to take child after work | 0.514 | 0.006 |
| 2. | 8 weeks | Partner too tired to take child after work | 0.898 | 0.004 |
| 3. | 8 weeks | Partner takes child after work, so mother is free | 0.708 | 0.006 |
| 4. | 8 weeks | Partner finds child hard to manage after work | 0.899 | 0.004 |
| 5. | 2 years 9 months | Partner finds it hard to cope with child after work | 0.303 | 0.016 |
| **Latent factor model fit indices** |  |  |  |  |
| Free parameters |  |  |  | 118 |
| Root Mean Square Error of Approximation (RMSEA) |  |  | 0.044 (95%CI |  |
|  |  |  | 0.043 to 0.045) |  |
| Comparative Fit Index (CFI) |  |  |  | 0.957 |
| Tucker-Lewis Index (TLI) |  |  |  | 0.950 |

**Figure 2.**
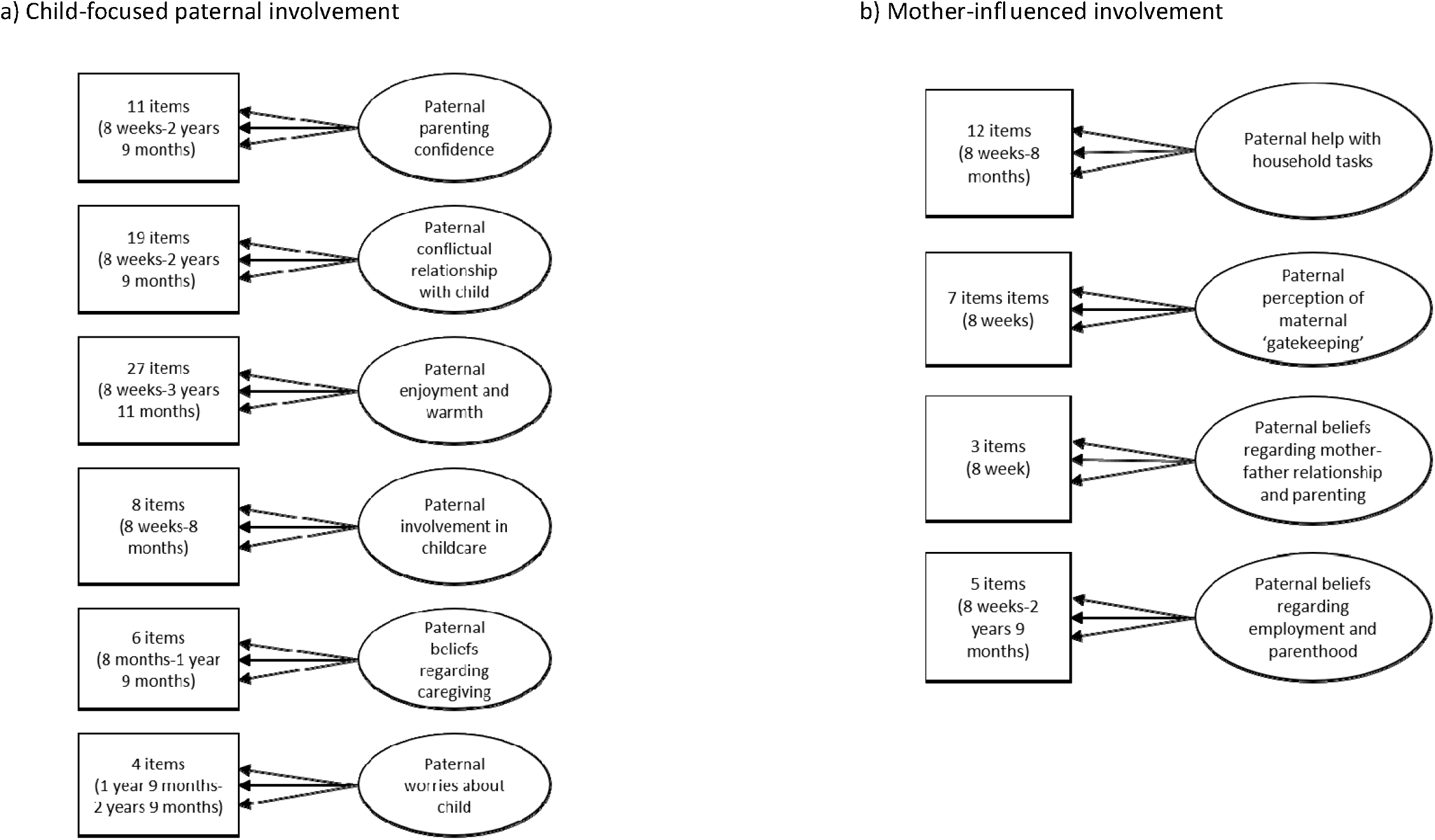
Hypothesised latent factor (CFA) measurement model representing specific factors capturing child-focused and mother-influenced dimensions of paternal involvement

**Figure 3.**
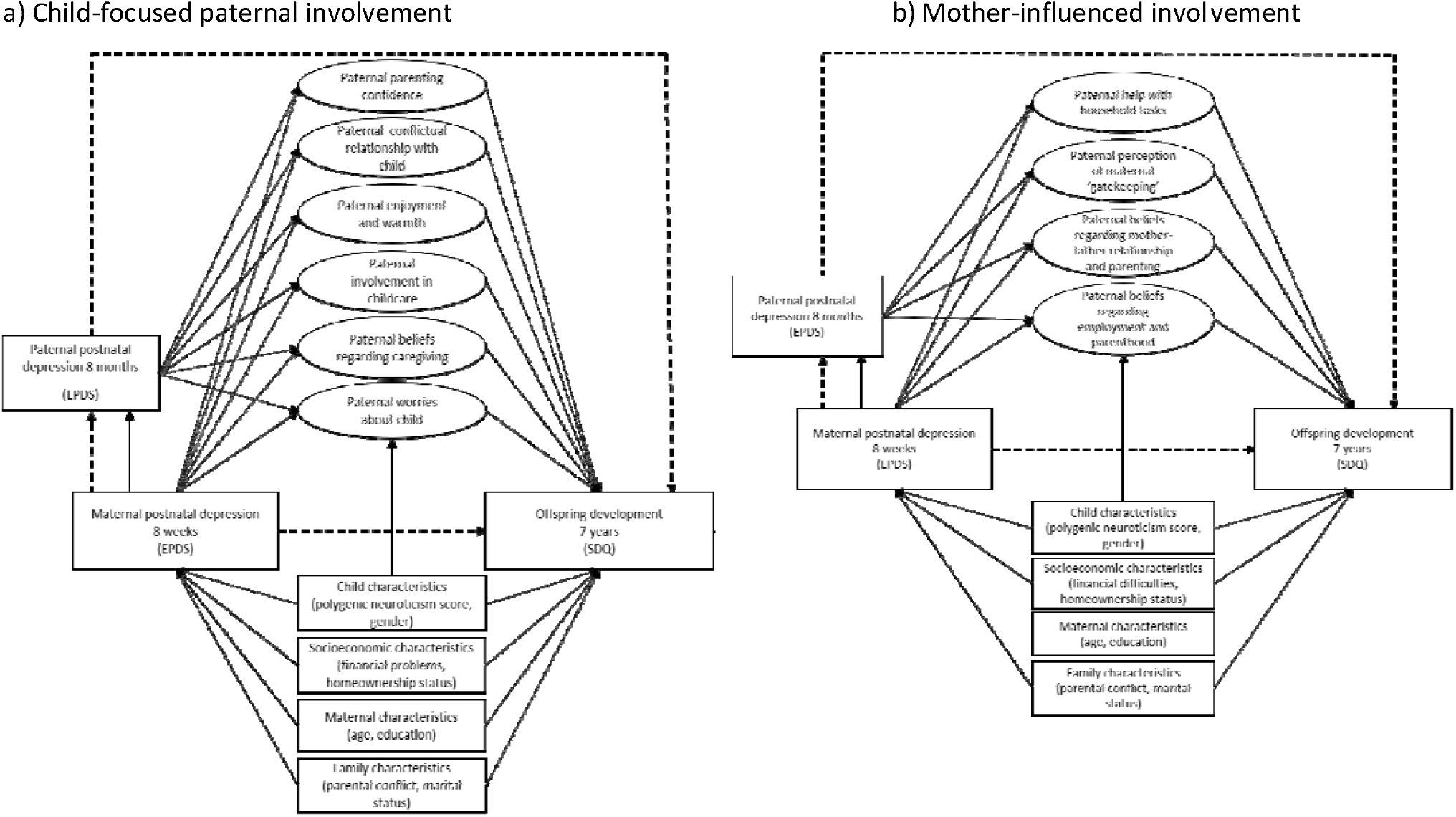
Structural equation mediation model estimating the direct effect of maternal postnatal depression on offspring development at age 7 years and the indirect effect through specific factors capturing child-focused and maternal-influenced dimensions of paternal involvement, adjusted for child PRS, gender, antenatal baseline confounders, and paternal PND as an intermediate confounder *Note:* Observed variables are represented by squares. Latent variables are represented by circles. Error terms covariances, correlations between the factors and individual items loading onto each specific factor are not represented to reduce figure complexity. Pathways that constitute total indirect effect are represented by bold lines; pathways that constitute direct effect are represented by dashed lines.

#### Potential baseline confounders: child polygenic score for neuroticism, socioeconomic, parental and family characteristics

Analyses were adjusted for child polygenic score (PGS) for neuroticism to account for possible child genetic correlations including evocative associations with parenting.^1^ However, it should be noted that polygenic scores capture only a small fraction of heritability, thus, they do not entirely capture possible genetic confounding.^51^ Genotyped data were available for 8,237 children in the ALSPAC cohort (full details in Methods S1, Supplementary). Disadvantaged socioeconomic status, marital status and conflict are also strong risk factors for maternal PND,^52^ less optimal paternal parenting practices,^53^ and adverse offspring emotional and behavioural development.^1^ In consequence, we adjusted for a range of antenatal prospectively measured potential confounding factors extracted from maternal questionnaires, including: highest maternal educational attainment (minimal education or none/compulsory secondary level [up to age 16 years; O-Level], non-compulsory secondary level [up to age 18 years; A-Level)/university level education]), maternal age in years, presence of financial difficulties (financial difficulties, no financial difficulties), homeownership status (owned/mortgaged, private/council [subsidised public housing] rented), marital status (married, never married) and a continuous score capturing parental conflict (higher scores representing higher levels of conflict). Given that our primary exposure of interest was maternal PND and lower response rates for fathers compared to mothers, as well as moderate to strong correlations between maternal and paternal education (r=0.6, p≤0.001) and age (r=0.7, p≤0.001), analyses were adjusted for mother-reported baseline confounders only to avoid introducing missing data.

#### Potential intermediate confounder: paternal postnatal depression

We also accounted for possible baseline antenatal confounders of the exposure-outcome, exposure-mediator and mediator-outcome associations by including them in the regression models to estimate each of these pathways.^44^ However, evidence suggests that maternal depression in pregnancy and during the postnatal period is associated with increased risk of paternal PND,^37,38^ which in turn influences paternal parenting,^39^ and offspring emotional and behavioural development,^40^ acting as a potential exposure induced intermediate confounder of the mediator-outcome association.^54,55^ Failure to account for intermediate confounders may result in biased inferences regarding direct and indirect (mediated) effects;^56-58^ thus, we accounted for paternal PND assessed using the Edinburgh Postnatal Depression Scale (EPDS)^45^ as a continuous score at 8 months postnatally (full details in Methods S1, Supplementary).

## Statistical analysis

### Latent factor models

Full details of latent factor models describing derivation of factors capturing child-focused and mother-influenced paternal involvement are presented in Methods S1, Supplementary. In summary, individual parenting items that were primarily theoretically relevant and had standardised loadings >0.15 were loaded onto the hypothesised parenting dimensions and modelled using Confirmatory Factor Analyses (CFA), a subset of SEM with a robust Weighted Least Square (WLSMV) estimator in M*plus* recommended to model both categorical and continuous data.^59^ The Root Mean Square Error of Approximation (RMSEA; >0.06) and Comparative Fit Index and Tucker-Lewis Index (CFI/TLI; >0.95) were used to evaluate the fit of the models.^60^ The chi-square test of overall fit is prone to model misspecification when sample size is large;^61^ thus, we gave preference to relative fit indices.

### Direct and mediated effects

Full details of the mediation model, including path-specific direct and indirect effects are described in Methods S1, Supplementary. In summary, we examined the extent to which the association between maternal PND (8 weeks) and offspring emotional and behavioural development (7 years) is explained (i.e., mediated) by child-focused (six latent factors; Figure 3a) and mother-influenced (four latent factors; Figure 3b) dimensions of paternal involvement using Structural Equation Modelling (SEM) in M*plus* v.8.3.^62^ Analyses of the mediation models were restricted to participants with complete data on exposure (maternal PND), outcome (offspring emotional and behavioural development), child PGS, baseline (socioeconomic, maternal and family characteristics) and intermediate (paternal PND) confounders (*n*=3,434).

First, we estimated the unadjusted models composed of exposure, outcome and mediators only. Second, we estimated models adjusted for child PGS and all antenatal baseline confounders (Model 1), and further adjusted for paternal PND (8 months) as a possible intermediate confounder of the mediator-outcome association (Model 2). Path-specific effects of interest representing pathways that constitute total indirect (bold lines) and direct (dashed lines) effects are described in Figure 3a-b and Methods S1, Supplementary. Indirect effects [95% CIs] were calculated using the product-of-coefficients method and bias-corrected (BC) bootstrapping (*n*=1,000 replications) to account for the non-normal distribution.^63^ Results from path analyses with continuous score (offspring total difficulties score), including indirect effects, are presented as unstandardised regression coefficients (hereafter referred to as *β*). Both results from Model 1 (adjusted for child PRS and antenatal baseline confounders) and Model 2 (further adjusted for paternal PND at 8 months as an intermediate confounder) are presented for comparison of estimates. However, preference is given to Model 2 under the assumption that Model 1 is biased without the inclusion of paternal PND.

### Missing data: multiple imputation

We conducted sensitivity analyses to examine the impact of missing data due to loss to follow-up on our findings. Full description of imputation methods is presented in Methods S1, Supplementary.

## Results

### Sample characteristics

Characteristics of the study sample and offspring total difficulties mean score at age 7 years by exposure status (maternal PND at 8 weeks) are presented in Results S1 (Table S1, Supplementary).

### Paternal involvement factors

CFA models to fit latent factors capturing child-focused and mother-influenced dimensions of paternal involvement suggested an adequate model fit (RMSEA: child-focused involvement: 0.022, 95%CI 0.021 to 0.022; mother-influenced involvement: 0.044, 95%CI 0.043 to 0.045; CFI/TLI: child-focused involvement: 0.904/0.899; mother-influenced involvement: 0.957/0.950; Tables 1-2) supporting further tests of structural paths (direct and mediated effects). Six factors capturing child-focused and four factors capturing mother-influenced dimensions of paternal involvement were derived (full details in Results S1), with associations between child-focused and mother-influenced factors presented in Results S1 (Table S2, Supplementary).

### Child-focused paternal involvement

#### Factor 1 Paternal parenting confidence

11 items relating to paternal feelings of confidence in the parenting role and perceptions of the ability to engage effectively in parenting behaviours, with higher factor scores representing higher levels of paternal parenting confidence.

#### Factor 2 Paternal conflictual relationship with child

19 items relating to conflict, harsh disciplining, irritation with the child and feelings of being overwhelmed, with higher factor scores signifying lower levels of conflictual parent-child relationship, irritation with the child and less harshness in paternal disciplining.

#### Factor 3 Paternal enjoyment and warmth

27 items relating to feelings of enjoyment, affection, love and warmth toward the child, with higher factor scores representing more paternal enjoyment, affection and warmth toward the child.

#### Factor 4 Paternal involvement in childcare

8 items describing frequency of paternal involvement in various aspects of childcare, with higher factor scores representing higher frequency of paternal involvement in childcare.

#### Factor 5 Paternal worries about child

4 items pertaining to paternal worries about the child, with higher factor scores representing lower levels of paternal worries about the child.

#### Factor 6 Paternal beliefs regarding caregiving

6 items relating to paternal beliefs regarding caregiving, including endorsement of structure (e.g., regularity and routines in infant care) and attunement (e.g., responsiveness to infant cues), with higher factor scores representing more paternal appreciation of structure and higher levels of attunement to child’s cues.

### Mother-focused paternal involvement

#### Factor 1 Paternal help with household tasks

12 items related to various aspects of paternal help with household tasks, with higher scores representing higher levels of paternal help with household tasks since the child was born.

#### Factor 2 Paternal perception of maternal ‘gatekeeping’

7 items relating to paternal perceptions of maternal beliefs and behaviours that encourage or hinder paternal involvement in childcare (i.e., maternal ‘gatekeeping’), with higher factor scores indicating higher levels of paternal perceptions of being supported and included in childcare by the mother (i.e., less maternal ‘gatekeeping’).

#### Factor 3 Paternal beliefs regarding mother-father relationship and parenting

3 items relating to paternal beliefs and perceptions regarding the impact that the birth of the child has had on the father-mother relationship, with higher factor scores representing paternal beliefs more concordant with positive changes in the nature of mother-father relationship following the birth of the child.

#### Factor 4 Paternal beliefs regarding employment and parenthood

5 items relating to paternal beliefs regarding maternal expectations around employment, childcare and paternal difficulties to manage childcare and employment, with higher scores indicating paternal beliefs concordant with less pressure to look after child after work and fewer difficulties with managing childcare and employment.

### Associations between maternal PND, child-focused and mother-influenced dimensions of paternal involvement and offspring emotional and behavioural development

Our pathways of interest were those between maternal PND and offspring emotional and behavioural development at age 7 years through child-focused and mother-influenced dimensions of paternal involvement. Specifically, we examined the pathways from maternal PND to paternal involvement (both direct and through paternal PND), and the pathways from paternal involvement to offspring emotional and behavioural development (Figure 3a-b represent pathways that constitute direct and total indirect effects) with full description of estimates for Model 1 provided in Results S1, Supplementary.

### Exposure (maternal PND) – mediator (paternal involvement) associations

To summarise, in Model 1 (adjusted for child PGS and antenatal baseline confounders; Table 3) maternal PND at 8 weeks was strongly associated with less paternal parenting confidence, more father-child conflict, less paternal enjoyment and warmth, more paternal worries about the child, higher levels of perceived maternal ‘gatekeeping’, more negative beliefs regarding the impact the birth of the child had on mother-father relationship, and paternal feelings of more pressure to look after the child after work and more struggles to manage childcare and employment.

**Table 3.** **Model 1:** Associations between maternal PND (8 weeks), offspring development (7 years) and dimensions of paternal involvement in the model adjusted for child PRS and all antenatal baseline confounders (n=3,434)

| Mediator: Paternal involvement | Exposure: Maternal PND |  | Outcome: Offspring development |  |
| --- | --- | --- | --- | --- |
|  | (8 weeks) |  | (7 years) |  |
|  | Fully adjusted model estimates <sup>a</sup> (n=3,434) |  |  |  |
|  | B [95% CI] | P-value | B [95% CI] | P-value |
| **Child-focused dimensions of paternal involvement** |  |  |  |  |
| *Paternal parenting confidence* | -0.049<br>[-0.061, -0.037] | ≤0.001 | -0.597<br>[-0.789, -0.404] | ≤0.001 |
| *Paternal conflictual relationship with child* | -0.053<br>[-0.063, -0.043] | ≤0.001 | -0.779<br>[-0.961, -0.597] | ≤0.001 |
| *Paternal enjoyment and warmth* | -0.032<br>[-0.042, -0.022] | ≤0.001 | -0.379<br>[-0.551, -0.206] | ≤0.001 |
| *Paternal involvement in childcare* | 0.007<br>[-0.003, 0.017] | 0.158 | -0.043<br>[-0.237, 0.151] | 0.664 |
| *Paternal beliefs regarding caregiving* | -0.009<br>[-0.234, 0.216] | 0.132 | -0.019<br>[-0.244, 0.206] | 0.872 |
| *Paternal worries about child* | -0.027<br>[-0.039, -0.015] | ≤0.001 | -0.277<br>[-0.479, -0.075] | 0.007 |
| **Mother-influenced dimensions of paternal involvement** |  |  |  |  |
| *Paternal help with household tasks* | 0.001<br>[-0.010, 0.011] | 0.932 | -0.016<br>[-0.186, 0.154] | 0.854 |
| *Paternal perception of maternal ‘gatekeeping’* | -0.040<br>[-0.054, -0.026] | ≤0.001 | -0.038<br>[-0.273, 0.197] | 0.749 |
| *Paternal beliefs regarding mother-father relationship* | -0.030<br>[-0.042, -0.018] | ≤0.001 | -0.040<br>[-0.234, 0.154] | 0.689 |
| *Paternal beliefs regarding employment and parenthood* | -0.031<br>[-0.041, -0.021] | ≤0.001 | -0.224<br>[-0.391, -0.057] | 0.008 |

Note:<sup>a</sup> Effect size are regression coefficients ( $\beta$ unstandardised) in the fully adjusted models: adjusted for child PGS, gender and antenatal baseline confounders (financial problems, homeownership status, maternal age and education, parental conflict, marital status).
Maternal PND: maternal postnatal depression; Child PGS: child polygenic score for neuroticism.

In contrast, in Model 2 (further adjusted for paternal PND at 8 months as an intermediate confounder; Table 4), maternal PND was strongly associated with more paternal enjoyment and warmth (β=0.055, 95% CI: 0.031, 0.078, p≤0.001) and direct involvement in childcare (β=0.029, 95% CI: 0.017, 0.041, p≤0.001), more paternal parenting confidence (β=0.029, 95% CI: 0.004, 0.054, p=0.027), less father-child conflict (β=0.034, 95% CI: 0.007, 0.061, p=0.016), more paternal help with household tasks (β=-0.009, 95% CI: -0.001, 0.019, p=0.070), and more paternal struggle to manage childcare and employment (β=-0.010, 95% CI: -0.019, -0.001, p=0.047), although 95% CIs were wide, that was not explained through paternal PND. Maternal PND was also strongly associated with paternal PND, which, in turn, was strongly associated with less paternal parenting confidence, less enjoyment and warmth, less involvement in childcare and appreciation of regular routine, more father-child conflict and more paternal worries about the child, less paternal help with household tasks, higher levels of perceived maternal ‘gatekeeping’, more negative feelings regarding the impact the birth of the child have had on mother-father relationship, and paternal feelings of more pressure to look after the child after work and more struggles to manage childcare and employment (Table 4).

**Table 4.** **Model 2:** Associations between maternal PND (8 weeks), offspring development (7 years), paternal PND (8 months), and dimensions of paternal involvement in the model adjusted for child PRS, gender, antenatal baseline confounders, and paternal PND as an intermediate confounder (n=3,434)

| Mediator: Paternal involvement | Exposure: Maternal PND |  | Outcome: Offspring development |  | Intermediate confounder: Paternal PND |  |
| --- | --- | --- | --- | --- | --- | --- |
|  | (8 weeks) |  | (7 years) |  | (8 months) |  |
|  | Fully adjusted model estimates <sup>a</sup> (n=3,434) |  |  |  |  |  |
|  | B [95% CI] | P-value | B [95% CI] | P-value | B [95% CI] | P-value |
| **Child-focused dimensions of paternal involvement** |  |  |  |  |  |  |
| *Paternal parenting confidence* | 0.029<br>[0.004, 0.054] | 0.027 | -0.309<br>[-0.732, 0.114] | 0.153 | -0.812<br>[-0.935, -0.689] | ≤0.001 |
| *Paternal conflictual relationship with child* | 0.034<br>[0.007, 0.061] | 0.016 | -0.546<br>[-0.998, -0.093] | 0.018 | -0.975<br>[-1.124, -0.826] | ≤0.001 |
| *Paternal enjoyment and warmth* | 0.055<br>[0.031, 0.078] | ≤0.001 | 0.072<br>[-0.316, 0.460] | 0.718 | -0.784<br>[-0.884, -0.684] | ≤0.001 |
| *Paternal involvement in childcare* | 0.029<br>[0.017, 0.041] | ≤0.001 | 0.099<br>[-0.110, 0.310] | 0.351 | -0.163<br>[-0.200, -0.128] | ≤0.001 |
| *Paternal beliefs regarding caregiving* | 0.004<br>[-0.010, 0.018] | 0.563 | 0.064<br>[-0.173, 0.301] | 0.595 | -0.102<br>[-0.141, -0.063] | ≤0.001 |
| *Paternal worries about child* | -0.011<br>[-0.023, 0.001] | 0.073 | -0.168<br>[-0.034, 0.370] | 0.104 | -0.117<br>[-0.150, -0.084] | ≤0.001 |
| *Maternal postnatal depression (8 weeks)* |  |  |  |  | 0.139<br>[0.115, 0.162] | ≤0.001 |
| *Offspring development (7 years)* |  |  |  |  | -0.410<br>[-0.533, 0.713] | 0.475 |
| **Mother-influenced dimensions of paternal involvement** |  |  |  |  |  |  |
| *Paternal help with household tasks* | 0.009<br>[-0.001, 0.019] | 0.070 | 0.039<br>[-0.135, 0.213] | 0.662 | -0.061<br>[-0.083, -0.039] | ≤0.001 |
| *Paternal perception of maternal ‘gatekeeping’* | -0.003<br>[-0.021, 0.015] | 0.771 | 0.216<br>[-0.109, 0.541] | 0.192 | -0.366<br>[-0.427, -0.305] | ≤0.001 |
| *Paternal beliefs regarding* | 0.007 | 0.508 | 0.295 | 0.100 | -0.416 | ≤0.001 |
| *mother-father relationship* | [-0.013, 0.027] |  | [-0.056, 0.646] |  | [-0.494, -0.338] |  |
| *Paternal beliefs regarding* | -0.010 | 0.047 | -0.097 | 0.300 | -0.169 | ≤0.001 |
| *employment and parenthood* | [-0.019, -0.001] |  | [-0.281, 0.087] |  | [-0.194, -0.143] |  |
| *Maternal postnatal depression* |  |  |  |  | 0.139 | ≤0.001 |
| *(8 weeks)* |  |  |  |  | [0.115, 0.162] |  |
| *Offspring development (7* |  |  |  |  | 0.337 | 0.019 |
| *years)* |  |  |  |  | [0.055, 0.619] |  |

*Note:*<sup>a</sup> Effect size are regression coefficients ( $\beta$ unstandardised) in the fully adjusted models: adjusted for child PGS, gender and antenatal baseline confounders (financial problems, homeownership status, maternal age and education, parental conflict, marital status).
Maternal PND: maternal postnatal depression; paternal PND: paternal postnatal depression; Child PGS: child polygenic score for neuroticism.

### Mediator (paternal involvement) – outcome (offspring emotional and behavioural development) associations

In Model 1, lower levels of paternal parenting confidence, warmth and enjoyment, higher levels of father-child conflict and worries about the child, and paternal difficulties to manage childcare and employment were associated with higher risk of offspring emotional and behavioural difficulties at age 7 years (Table 3). In Model 2, however, the only child-focused dimension of paternal involvement, higher levels of father-child conflict, was strongly associated with higher risk of offspring emotional and behavioural difficulties at age 7 years (β=-0.546, 95% CI: -0.998, -0.093, p=0.018; Table 4), with no evidence for associations between any of the mother-influenced dimensions of paternal involvement and offspring emotional and behavioural development.

### Direct and mediated effects

Full description of estimates for direct and mediated effects in Model 1 is presented in Results S1, Supplementary. To summarise, in Model 1 (Table 5), there was evidence of a total indirect effect from maternal PND to offspring emotional and behavioural development at age 7 years through the combination of all child-focused, but not mother-influenced, dimensions of paternal involvement. There was strong evidence of total and direct effects from maternal PND to offspring development in models capturing both child-focused and mother-influenced paternal involvement. There was evidence for specific indirect effects through paternal parenting confidence, paternal conflictual relationship with the child, paternal enjoyment and warmth, paternal worries about the child, and paternal difficulties to manage childcare and employment (although 95% CIs were wide).

**Table 5.**
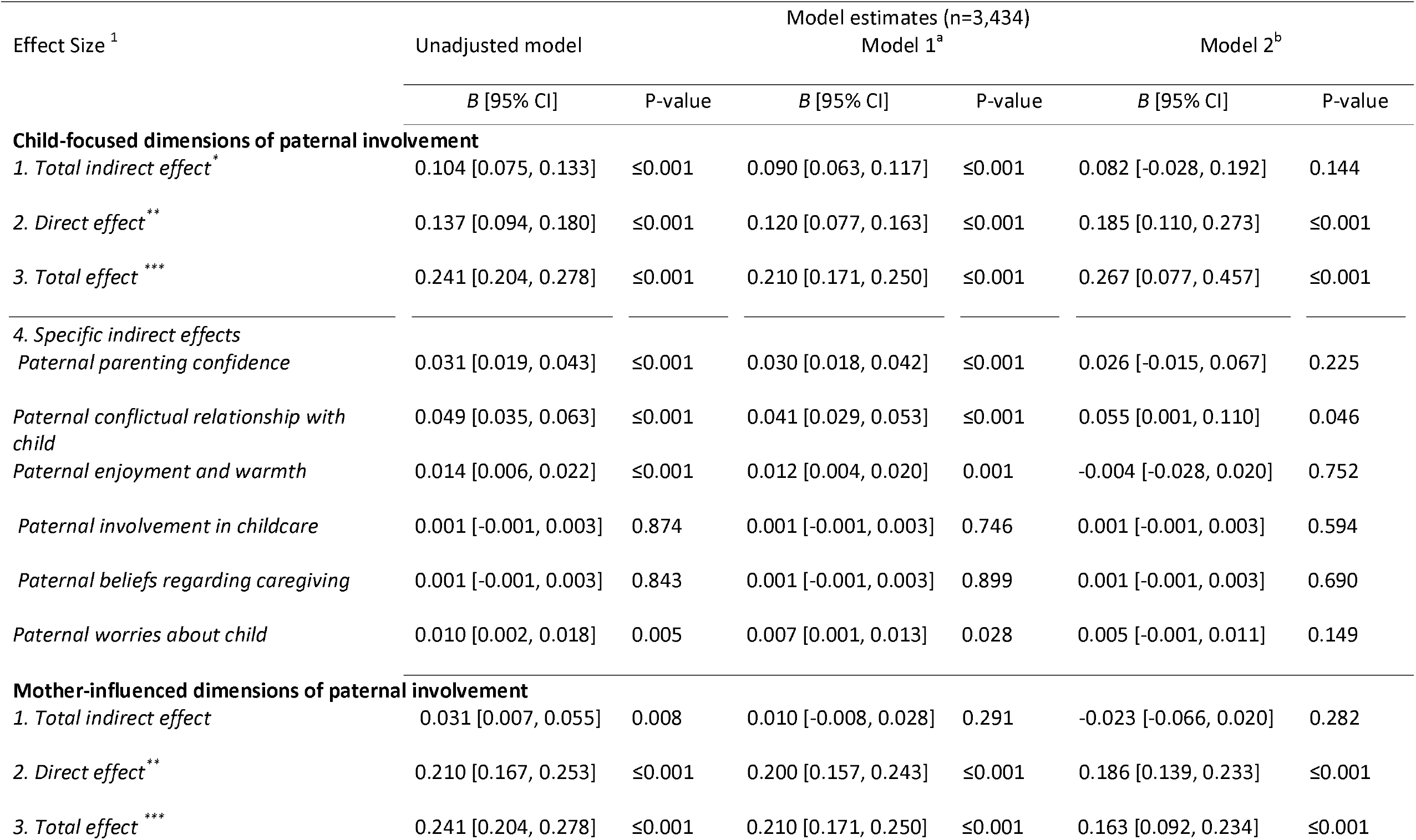
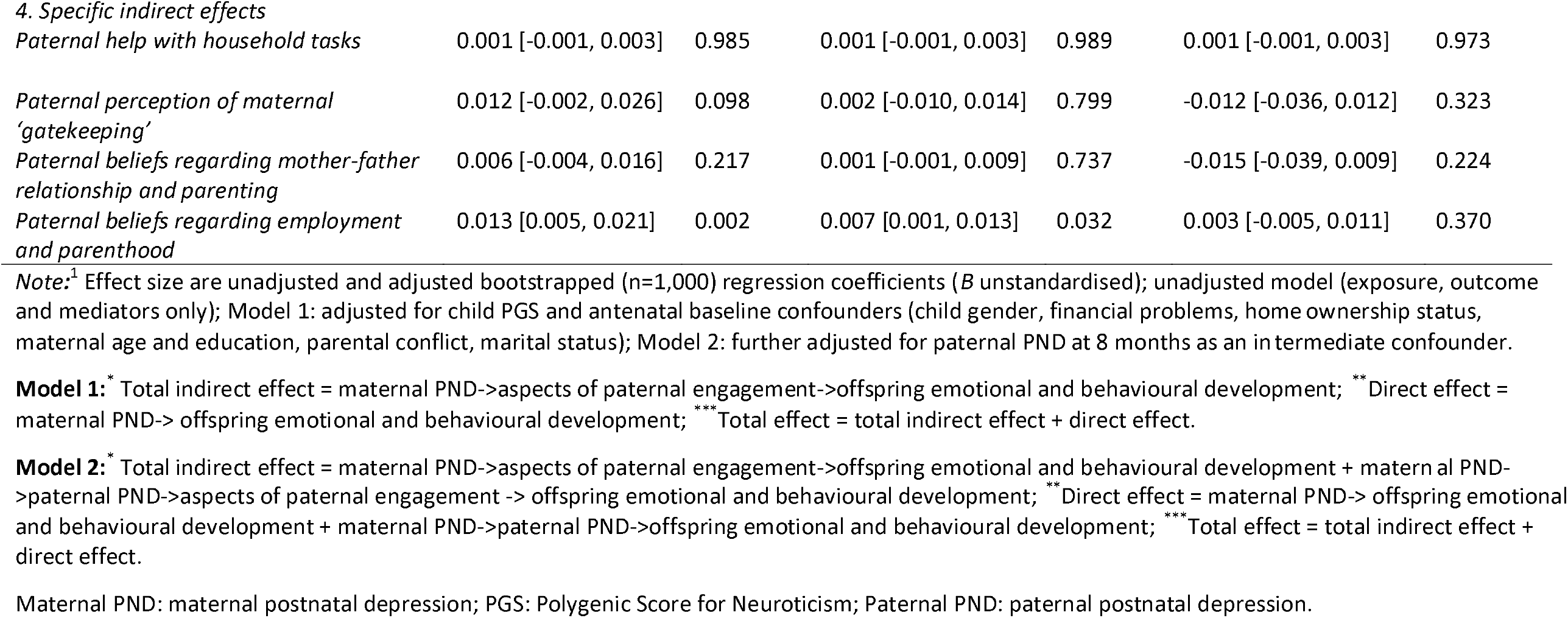
Estimates of direct and mediated effects in the unadjusted model and models adjusted for child PRS, gender, antenatal baseline confounders, and paternal PND as an intermediate confounder in complete sample (n=3,434)

| Effect Size <sup>1</sup> | Model estimates (n=3,434) |  |  |  |  |  |
| --- | --- | --- | --- | --- | --- | --- |
|  | Unadjusted model |  | Model 1<sup>a</sup> |  | Model 2<sup>b</sup> |  |
|  | *B* [95% CI] | P-value | *B* [95% CI] | P-value | *B* [95% CI] | P-value |
| **Child-focused dimensions of paternal involvement** |  |  |  |  |  |  |
| 1. Total indirect effect<sup>*</sup> | 0.104 [0.075, 0.133] | ≤0.001 | 0.090 [0.063, 0.117] | ≤0.001 | 0.082 [-0.028, 0.192] | 0.144 |
| 2. Direct effect<sup>**</sup> | 0.137 [0.094, 0.180] | ≤0.001 | 0.120 [0.077, 0.163] | ≤0.001 | 0.185 [0.110, 0.273] | ≤0.001 |
| 3. Total effect<sup>***</sup> | 0.241 [0.204, 0.278] | ≤0.001 | 0.210 [0.171, 0.250] | ≤0.001 | 0.267 [0.077, 0.457] | ≤0.001 |
| **4. Specific indirect effects** |  |  |  |  |  |  |
| Paternal parenting confidence | 0.031 [0.019, 0.043] | ≤0.001 | 0.030 [0.018, 0.042] | ≤0.001 | 0.026 [-0.015, 0.067] | 0.225 |
| Paternal conflictual relationship with child | 0.049 [0.035, 0.063] | ≤0.001 | 0.041 [0.029, 0.053] | ≤0.001 | 0.055 [0.001, 0.110] | 0.046 |
| Paternal enjoyment and warmth | 0.014 [0.006, 0.022] | ≤0.001 | 0.012 [0.004, 0.020] | 0.001 | -0.004 [-0.028, 0.020] | 0.752 |
| Paternal involvement in childcare | 0.001 [-0.001, 0.003] | 0.874 | 0.001 [-0.001, 0.003] | 0.746 | 0.001 [-0.001, 0.003] | 0.594 |
| Paternal beliefs regarding caregiving | 0.001 [-0.001, 0.003] | 0.843 | 0.001 [-0.001, 0.003] | 0.899 | 0.001 [-0.001, 0.003] | 0.690 |
| Paternal worries about child | 0.010 [0.002, 0.018] | 0.005 | 0.007 [0.001, 0.013] | 0.028 | 0.005 [-0.001, 0.011] | 0.149 |
| **Mother-influenced dimensions of paternal involvement** |  |  |  |  |  |  |
| 1. Total indirect effect | 0.031 [0.007, 0.055] | 0.008 | 0.010 [-0.008, 0.028] | 0.291 | -0.023 [-0.066, 0.020] | 0.282 |
| 2. Direct effect<sup>**</sup> | 0.210 [0.167, 0.253] | ≤0.001 | 0.200 [0.157, 0.243] | ≤0.001 | 0.186 [0.139, 0.233] | ≤0.001 |
| 3. Total effect<sup>***</sup> | 0.241 [0.204, 0.278] | ≤0.001 | 0.210 [0.171, 0.250] | ≤0.001 | 0.163 [0.092, 0.234] | ≤0.001 |
| 4. Specific indirect effects |  |  |  |  |  |  |
| *Paternal help with household tasks* | 0.001 [-0.001, 0.003] | 0.985 | 0.001 [-0.001, 0.003] | 0.989 | 0.001 [-0.001, 0.003] | 0.973 |
| *Paternal perception of maternal 'gatekeeping'* | 0.012 [-0.002, 0.026] | 0.098 | 0.002 [-0.010, 0.014] | 0.799 | -0.012 [-0.036, 0.012] | 0.323 |
| *Paternal beliefs regarding mother-father relationship and parenting* | 0.006 [-0.004, 0.016] | 0.217 | 0.001 [-0.001, 0.009] | 0.737 | -0.015 [-0.039, 0.009] | 0.224 |
| *Paternal beliefs regarding employment and parenthood* | 0.013 [0.005, 0.021] | 0.002 | 0.007 [0.001, 0.013] | 0.032 | 0.003 [-0.005, 0.011] | 0.370 |

*Note:*<sup>1</sup> Effect size are unadjusted and adjusted bootstrapped (n=1,000) regression coefficients (*B* unstandardised); unadjusted model (exposure, outcome and mediators only); Model 1: adjusted for child PGS and antenatal baseline confounders (child gender, financial problems, homeownership status, maternal age and education, parental conflict, marital status); Model 2: further adjusted for paternal PND at 8 months as an intermediate confounder.
**Model 1:**\* Total indirect effect = maternal PND->aspects of paternal engagement->offspring emotional and behavioural development; \*\* Direct effect = maternal PND-> offspring emotional and behavioural development; \*\*\* Total effect = total indirect effect + direct effect.
**Model 2:**\* Total indirect effect = maternal PND->aspects of paternal engagement->offspring emotional and behavioural development + maternal PND->paternal PND->aspects of paternal engagement -> offspring emotional and behavioural development; \*\* Direct effect = maternal PND-> offspring emotional and behavioural development + maternal PND->paternal PND->offspring emotional and behavioural development; \*\*\* Total effect = total indirect effect + direct effect.
Maternal PND: maternal postnatal depression; PGS: Polygenic Score for Neuroticism; Paternal PND: paternal postnatal depression.

In contrast, in Model 2 (Table 5), there was no evidence of total indirect effect from maternal PND to offspring development at 7 years through the combination of all factors capturing child-focused (β=0.082, 95%CIs: -0.028, 0.192, p=0.144) and mother-influenced (β=-0.023, 95%CIs: -0.066, 0.020, p=0.282) dimensions of paternal involvement. Similar to Model 1, there was strong evidence of total and direct effects from maternal PND to offspring development in models capturing both child-focused (total effect: β=0.267, 95%CIs: 0.077, 0.457, p≤0.001; direct effect: β=0.185, 95%CIs: 0.110, 0.273, p≤0.001) and mother-influenced (total effect: β=0.163, 95%CIs: 0.092, 0.234, p≤0.001; direct effect: β=0.186, 95%CIs: 0.139, 0.233, p≤0.001) paternal involvement. However, unlike Model 1, there was only some evidence for specific indirect effect through paternal conflictual relationship with the child (β=0.055, 95%CIs: 0.001, 0.110, p=0.046), although the 95% CIs were wide, but not other child-focused dimensions of paternal involvement. There was no evidence for specific indirect effects through any of the mother-influenced dimensions of paternal engagement.

## Discussion

### Main findings

In this population-based cohort study, we modelled various dimensions of child-focused and mother-influenced dimensions of paternal involvement during early childhood and estimated the extent to which they explain the association between maternal PND and offspring emotional and behavioural difficulties at age 7 years. One of the strengths of our study is the inclusion of paternal PND as a possible exposure-induced intermediate confounder of the mediator-outcome association (see Figure 1 for explanation of conceptual pathways). It has been extensively argued that failure to account for potential intermediate confounders may bias inferences regarding direct and indirect (mediated) effects.^56-58^

In our models adjusted for child PGS and baseline confounders only, maternal PND was strongly associated with *lower scores* on most child-focused and mother-influenced dimensions of paternal involvement, which explained (mediated) a proportion of the association between maternal PND and offspring emotional and behavioural difficulties. However, accounting for paternal PND *reversed* associations between maternal PND and both child-focused and mother-influenced dimensions of paternal involvement. Specifically, once paternal PND was accounted for, maternal PND was associated with *higher scores* on child-focused and mother-influenced paternal involvement, including higher levels of paternal parenting confidence, enjoyment and warmth toward the child, less father-child conflict, and more paternal involvement in childcare and more help with household tasks. This finding suggests that fathers’ own mental health is likely to influence paternal involvement in the context of maternal PND, with fathers who are not depressed developing ‘compensatory’ parenting in response to their partner’s depression.^6,13,14^ In contrast, the ‘spillover’ hypothesis, wherein higher levels of maternal depressive symptoms are associated with lower father involvement,^6^ was only evident in the model that did not account for paternal PND. Further research is warranted to examine a possible protective role of child-focused and mother-influenced dimensions of paternal involvement in the context of maternal and paternal PND via a moderation hypothesis (i.e., whether the risk of adverse offspring development varies according to levels of paternal involvement).

Our findings indicated that maternal PND was still associated with more paternal struggles to manage employment and childcare responsibilities once paternal PND was accounted for, suggesting that these dimensions of paternal involvement are negatively associated with maternal PND irrespective of paternal PND. This finding is consistent with existing sociological literature emphasising paternal struggles to maintain ‘work-life balance’,^29,64^ and a fractious relationship between paid employment and family activities,^65^ often placed in opposition to each other.

Amongst all dimensions of paternal involvement, only conflictual father-child relationship emerged as a risk factor for adverse offspring development, as well as an explanatory mechanism, albeit the effect size was small, in the association between maternal PND and offspring development irrespective of paternal mental health. There was no evidence for the mediating role of any other child-focused and mother-influenced dimensions of paternal involvement. It has previously been argued that conflictual father-child relationship may undermine children’s sense of emotional security increasing susceptibility for emotional and behavioural problems even in the absence of paternal PND.^31^ Paternal PND has also been linked to offspring emotional and behavioural difficulties and father-child conflict,^66^ which, in turn, mediates the association between paternal PND and adverse offspring development.^67^ Our findings suggest that father-child conflict may also be a mechanism of risk transmission in the context of maternal PND even if the father is not depressed.

Strong evidence emerged for a direct association between maternal PND and risk of offspring adverse emotional and behavioural development in both models examining child-focused and mother-influenced dimensions of paternal involvement. These findings are consistent with previous research suggesting that paternal non-involvement, modelled as a sum-score, explained a small proportion of the association between maternal PND and offspring development.^68^ It may be that other pathways of transmission from maternal PND to offspring development, for instance quality of mother-child relationship and maternal parenting, may be more important in this context.^69^ Consistent with accumulating epidemiological evidence, maternal PND was associated with increased risk of paternal PND,^38,70,71^ which, in turn, was strongly associated with increased risk of adverse offspring emotional and behavioural development.^40,72^ In line with previous research, paternal PND was also strongly associated with reduced child-focused and mother-influenced paternal involvement.^39^

### Strengths and limitations

Strengths of this study include a longitudinal design, large community-based sample and availability of rich repeated self-reported measures of paternal parenting that enabled us to model several dimensions of child-focused and mother-influenced dimensions of paternal involvement across early childhood, as well as to examine their possible explanatory (i.e., mediating) role in the association between maternal PND and offspring emotional and behavioural development in mediation models adjusted for a range of confounders, including paternal PND and child PGS. To the best of our knowledge, no previous studies have examined dimensions of child-focused and mother-influenced involvement as possible transmission pathways in the association between maternal PND and offspring development while accounting for paternal PND.

Paternal involvement is embedded within complex family systems of intertwined relationships and their impact on the nature of parent-child relationship.^9,19,24,73^ Our conceptualisation of paternal involvement was guided by developmental and sociological literature explicitly acknowledging the complexity of paternal roles and the importance of affective and cognitive, not just behavioural,^31^ dimensions, as well as the negotiated nature of paternal involvement shaped by significant relationships.^24,74-76^ Although this perspective renders conceptual and statistical examination of paternal involvement more difficult,^7,17^ it answers the call for multidimensional conceptualisation of father involvement,^31^ enabling a more nuanced understanding of the variety of forms in which paternal involvement occurs and their individual contributions to child development.

Associations among parental mental health, parenting and offspring development are complex and bidirectional.^34,35^ Our findings suggest that maternal PND is associated with paternal PND, which, in turn, is associated with reduced child-focused and mother-influenced dimensions of paternal involvement. However, in line with transactional developmental models,^77^ children with more difficult emotional and behavioural responses may influence maternal and paternal PND, as well as paternal involvement.^78,79^ Addressing possible bidirectionality^80^ fell outside the scope of the present study. We attempted to account for possible evocative effects and possible shared genetic liability for emotional difficulties in parents and offspring by including a child genetic score for neuroticism, which explained a small proportion in these traits. However, polygenic scores capture a small fraction of trait heritability and, thus, of the confounding due to genetic factors unlikely to reflect true genetic effects.^35^ Shared genetic variance in depression and parenting not captured by the score may still confound the associations between parental PND, paternal involvement and offspring development.

Another limitation is non-independence of measurement. Maternal PND and offspring development were both reported by mothers. Evidence suggests that depressed mothers may inaccurately report their children’s emotional and behavioural problems,^81,82^ potentially overestimating the direct effect. The complexity of the model precluded us from examining offspring outcome as a multi-informant latent factor (i.e., rated by parents, clinicians and teachers)^83^ to address this limitation. However, given the strength of the direct association between maternal PND and offspring emotional and behavioural difficulties modelled as the SDQ total problems score, any multi-informant latent factor would unlikely change this finding. Modelling SDQ as total difficulties score is a well-validated approach widely adopted across development studies, including those utilising ALSPAC data.^49^ In addition, paternal involvement measures were father-reported, reducing the possibility of overestimating mediated effects (i.e., the associations between maternal and paternal PND, paternal involvement and offspring development) due to shared variance bias. The importance of independent data in examining mediating pathways between maternal PND and offspring development has been strongly articulated.^84^ We also modelled paternal involvement items across several time points in childhood, arguably painting a more fine-grained picture of paternal involvement compared to any one-off assessment through direct/independent observations. However, our findings do not imply a ‘unique’ role for paternal child-focused and mother-influenced involvement over and above maternal involvement, which we did not account for due to lack of comparative measures.

Inter-parental conflict, as a measure of family functioning, is an important factor in the context of parental depression, parenting and offspring development.^85^ Analyses were adjusted for antenatal levels of inter-parental conflict; however, we did not examine its role in the association between maternal PND and offspring development at later time points.^86^ Examination of interrelated and, most likely, bi-directional associations among parental PND, inter-parental conflict, paternal involvement and offspring development was beyond the scope of this study, warranting further research to disentangle these complex pathways.

### Implications of the research and conclusions

Our findings emphasise the importance of the widely articulated, but rarely adopted, clinical stance to consider both maternal and paternal mental health when one parent presents with depression. Recognising and addressing paternal PND during the postnatal period will not only support fathers but also improve their ability to support their partners and children. The findings of this study also highlight the importance of involving fathers in the treatment of maternal depression through partner-assisted therapies, such as interpersonal counselling,^87,88^ as well as interventions that address the importance of paternal engagement and caregiving directly via education and, for instance, video-feedback.^89-91^ Given the role that father-child conflict plays in transmitting the negative effect of maternal PND on offspring development even in the absence of paternal PND, and should these effects be found causal in future research, targeted interventions to reduce father-child conflict may contribute to reducing intergenerational transmission of mental health risks in offspring of depressed mothers. Nonetheless, it is important to note that maternal PND was strongly associated with adverse offspring development even in the context of paternal involvement, suggesting that addressing maternal PND remains a key factor in reducing adverse offspring outcomes.

The mother-child relationship has been a cornerstone of developmental research, often at the expense of the wider family system perspective, which explicitly acknowledges the father-child relationship as a crucial sub-system, integrated and influential in the context of higher-order family system.^92,93^ This has important implications for policy and practice that should encourage and facilitate father involvement in interventions designed to improve maternal and offspring outcomes, crucially without losing sight of the importance of treatment for mothers. Given the high prevalence of maternal PND and the substantial health and societal burden it potentially inflicts if left unsupported,^94^ policies that facilitate and advance father involvement in the context of maternal PND in particular are of great importance for family health and well-being.

## Data Availability

If you have any questions about accessing data, please.

## Ethical standards

Informed consent for the use of data collected via questionnaires and clinics was obtained from participants following the recommendations of the ALSPAC Ethics and Law Committee at the time.

## Acknowledgements

We are extremely grateful to all the families who took part in this study, the midwives for their help in recruiting them, and the whole ALSPAC team, which includes interviewers, computer and laboratory technicians, clerical workers, research scientists, volunteers, managers, receptionists and nurses. Professor Marinus H Van Ijzendoorn has generously provided insightful review and comments on the earlier versions of this manuscript.

## References

1. Stein, A., Pearson, R.M., Goodman, S.H., Rapa, E., Rahman, A., McCallum, M., Howard, L.M., & Pariante, C.M. (2014). Effects of perinatal mental disorders on the fetus and child. The Lancet, 384, 1800–1819.

2. Mezulis, A. H., Hyde, J. S., & Clark, R. (2004). Father involvement moderates the effect of maternal depression during a child’s infancy on child behavior problems in kindergarten. Journal of Family Psychology, 18, 575–588.

3. Sanger, C., Iles, J. E., Andrew, C. S., & Ramchandani P.G. (2015). Associations between postnatal maternal depression and psychological outcomes in adolescent offspring: a systematic review. Archives of Women Mental Health, 18, 147–162.

4. Murray, L., Halligan, S. L., & Cooper, P. (2010). Effects of postnatal depression on mother-infant interactions, and child development. In: Handbook of infant development, eds. Malden, UK: Wiley-Blackwell.

5. Giallo, R., Cooklin, A., Wade, C., D’Esposito, F., & Nicholson, J. M. (2014). Maternal postnatal mental health and later emotional–behavioural development of children: the mediating role of parenting behaviour. Child: Care, Health and Development, 40, 327–336.

6. Goodman, S. H., Lusby, C. M., Thompson, K., Newport, D. J., & Stowe, Z. N. (2014). Maternal depression in association with fathers’ involvement with their infants: spillover or compensation/buffering? Infant Mental Health Journal, 35, 495–508.

7. Tamis-LeMonda, C. S., & Cabrera, N. J. (Eds.) (2002). Handbook of father involvement: Multidisciplinary perspectives. Mahwah, NJ: Erlbaum.

8. Carlson, M. J. (2006). Family structure, father involvement, and adolescent behavioral outcomes. Journal of Marriage and Family, 68, 137–154.

9. Lamb, M. E. (2010). How do fathers influence children’s development? Let me count the ways. In M.E. Lamb (Eds.), The role of the father in child development, pp. 1–26. Hoboken, NJ: Wiley.

10. Sarkadi, A., Kristiansson, R., Oberklaid, F., & Bremberg, S. (2008). Fathers’ involvement and children’s developmental outcomes: a systematic review of longitudinal studies. Acta Paediatrica, 97, 153–158.

11. Masten, A. S., & Monn, A. R. (2015). Child and family resilience: A call for integrated science, practice, and professional training. Family Relations, 64, 5–21.

12. Patterson, J. M. (2002). Understanding family resilience. Journal of Clinical Psychology, 58, 233–246.

13. Edhborg, M., Lundh, W., Seimyr, L., & Widström, A. M. (2003). The parent-child relationship in the context of maternal depressive mood. Archives of Women’s Mental Health, 6, 211–216.

14. Hossain, Z., Field, T., Gonzalez, J., Malphurs, J., Valle, C. D., & Pickens, J. (1994). Infants of “depressed” mothers interact better with their nondepressed fathers. Infant Mental Health Journal, 15, 348–357.

15. Vakrat, A., Apter-Levy, Y., & Feldman, R. (2018). Sensitive fathering buffers the effects of chronic maternal depression on child psychopathology. Child Psychiatry & Human Development, 49, 779–785.

16. Carro, M. G., Grant, K. E., Gotlib, I. H., & Compas, B. E. (1993). Postpartum depression and child development: An investigation of mothers and fathers as sources of risk and resilience. Development and Psychopathology, 5, 567–579.

17. Chabrol, H., Bron, N., & Le Camus, J. (1996). Mother-infant and father-infant interactions in postpartum depression. Infant Behavior and Development, 19, 149–152.

18. Paulson, J. F., Dauber, S. E., & Leiferman, J. A. (2011). Parental depression, relationship quality, and nonresident father involvement with their infants. Journal of Family Issues, 32, 528–549.

19. Lamb, M. E., & Lewis, C. (2004). The development and significance of father–child relationships in two-parent families. In M. E. Lamb (Eds.), The role of the father in child development (4th ed., pp. 272–306). Hoboken, NJ: Wile

20. Tamis-LeMonda, C. S., Shannon, J. D., Cabrera, N. J., & Lamb, M. E. (2004). Fathers and mothers at play with their 2-and 3-year-olds: Contributions to language and cognitive development. Child Development, 75, 1806–1820.

21. Dermott, E. (2003). The ‘intimate father’: Defining paternal involvement. Sociological Research Online, 8, 28–38.

22. Parke, R. D., & Cookston, J. T. (2019). Fathers and family. In M. H. Bornstein (Eds.), Handbook of parenting. Vol.3. Being and becoming a parent (3rd ed., pp.64–136). New York: Routlege.

23. Pleck, J. H. (2010). Paternal involvement: Revised conceptualisation and theoretical linkages with child outcomes. In M. Lamb (Eds.), The role of the father in child development (pp. 58–93). Hoboken, NJ: Wiley.

24. Marsiglio, W., Day, R. D., & Lamb, M. E. (2000). Exploring fatherhood diversity: Implications for conceptualizing father involvement. Marriage & Family Review, 29, 269–293.

25. Miller, T. (2018). Paternal and maternal gatekeeping? Choreographing care. Sociologica, 12, 25–35.

26. Pleck, J. H. (2012). Integrating father involvement in parenting research. Parenting, 12, 243–253.

27. Schoppe-Sullivan, S. J., & Altenburger, L. E. (2019). In M. H. Bornstein (Eds.), Handbook of parenting. Vol.3. Being and becoming a parent (3rd ed., pp.64–136). New York: Routlege.

28. Lamb, M. E. (2013). Father-child relationships. In C. S. Tamis-LeMonda, & N. J. Cabrera, (Eds.) Handbook of father involvement: Multidisciplinary perspectives (pp.119–134). Mahwah, NJ: Erlbaum.

29. Miller, T. (2010). Making sense of fatherhood: Gender, caring and work. Cambridge University Press.

30. Cabrera, N. J., Fitzgerald, H. E., Bradley, R. H., & Roggman, L. (2014). The ecology of father-child relationships: An expanded model. Journal of Family Theory & Review, 6(4), 336–354.

31. Palkovitz, R. (2019). Expanding our focus from father involvement to father–child relationship quality. Journal of Family Theory & Review, 11(4), 576–591.

32. Schoppe-Sullivan, S. J., McBride, B. A., & Ho, M. H. R. (2004). Unidimensional versus multidimensional perspectives on father involvement. Fathering: A Journal of Theory, Research & Practice about Men as Fathers, 2, 147–164.

33. Bornstein, M. H. (2015). Children’s Parents. In: Ecological settings and processes in developmental systems. Handbook of child psychology and developmental science. Hoboken, NJ: Wiley.

34. Mills-Koonce, W. R., Propper, C. B., Gariepy, J. L, Blair, C., Garrett-Peters, P., & Cox M. J. (2007). Bidirectional genetic and environmental influences on mother and child behavior: the family system as the unit of analyses. Developmental Psychopathology, 19, 1073–1087.

35. Avinun, R., & Knafo, A. (2014). Parenting as a reaction evoked by children’s genotype: a meta-analysis of children-as-twins studies. Personality and Social Psychology Review, 18, 87–102.

36. Pettit, G. S., & Arsiwalla, D. D. (2008). Commentary on special section on “bidirectional parent– child relationships”: The continuing evolution of dynamic, transactional models of parenting and youth behavior problems. Journal of Abnormal Child Psychology, 36, 711–718.

37. Goodman, J. H. (2004). Paternal postpartum depression, its relationship to maternal postpartum depression, and implications for family health. Journal of Advanced Nursing, 45, 26–35.

38. Fredriksen, E., Von Soest, T., Smith, L., & Moe, V. (2019). Depressive symptom contagion in the transition to parenthood: Interparental processes and the role of partner-related attachment. Journal of Abnormal Psychology, 128, 397–403.

39. Wilson, S., & Durbin, C. E. (2010). Effects of paternal depression on fathers’ parenting behaviors: A meta-analytic review. Clinical Psychology Review, 30, 167–180.

40. Ramchandani, P., Stein, A., Evans, J., O’Connor, T. G., & ALSPAC Study Team. (2005). Paternal depression in the postnatal period and child development: a prospective population study. The Lancet, 365, 2201–2205.

41. Kaminski, J. W., Valle, L. A., Filene, J. H., & Boyle, C. L. (2008). A meta-analytic review of components associated with parent training program effectiveness. Journal of Abnormal Child Psychology, 36, 567–589.

42. Boyd, A., Golding, J., Macleod, J., Lawlor, D.A., Fraser, A., Henderson, J., Molloy, L., Ness, A., Ring, S., & Davey Smith, G. (2013). Cohort Profile: The ‘Children of the 90s’: the index offspring of the Avon Longitudinal Study of Parents and Children (ALSPAC). International Journal of Epidemiology, 42, 111–127.

43. Fraser, A., Macdonald-Wallis, C., Tilling, K., Boyd, A., Golding, J., Davey Smith, G., Henderson, J., Macleod, J., Molloy, L., Ness, A., Rong, S., Nelson, S.M., & Lawlor, D.A. (2013). Cohort profile: The Avon Longitudinal Study of Parents and Children: ALSPAC mothers cohort. International Journal of Epidemiology, 42, 97–110.

44. Northstone, K., Lewcock, M., Groom, A., Boyd, A., Macleod, J., Timpson, N.J., & Wells, N. (2019). The Avon Longitudinal Study of Parents and Children (ALSPAC): an update on the enrolled sample of index children in 2019. Wellcome Open Research, 4:51. DOI: https://doi.org/10.12688/wellcomeopenres.15132.1.

45. Cox, J. L., Holden, J. M., & Sagovsky, R. (1987). Detection of postnatal depression: development of the 10-item Edinburgh Postnatal Depression Scale. British Journal of Psychiatry, 150, 782–786.

46. Pearson, R. M., Evans, J., Kounali, D., Lewis, G., Heron, J., Ramchandani, P. G., O’Connor, T.G., & Stein, A. (2013). Maternal depression during pregnancy and the postnatal period: risks and possible mechanisms for offspring depression at age 18 years. JAMA Psychiatry, 70, 1312–1319.

47. Goodman, R. S. (1997). Strengths and Difficulties Questionnaire: a research note. Journal of Child Psychology and Psychiatry, 38, 581–586.

48. Goodman, R. (2001). Psychometric properties of the strengths and difficulties questionnaire. Journal of American Academy of Child and Adolescent Psychiatry, 40, 1337–1345.

49. Gutierrez-Galve, L., Stein, A., Hanington, L., Heron, J., & Ramchandani, P. (2015). Paternal depression in the postnatal period and child development: mediators and moderators. Pediatrics, 135(2), e339–e347.

50. Dermott, E. (2014). Intimate fatherhood: A sociological analysis. Routledge.

51. Pingault, J. B., Rijsdijk, F., Schoeler, T., Choi, S. W., Selzam, S., Krapohl, E., O’Reilly, P. F., & Dudbridge, F. (2021). Genetic sensitivity analysis: adjusting for genetic confounding in epidemiological associations. PLoS genetics, 17(6), e1009590.

52. Milgrom, J., Gemmill, A. W., Bilszta, J. L., Hayes, B., Barnett, B., Brooks, J., Ericksen, J., Ellwood, D., & Buist, A. (2008). Antenatal risk factors for postnatal depression: a large prospective study. Journal of Affective Disorders, 108, 147–157.

53. Leigh, B., & Milgrom, J. (2008). Risk factors for antenatal depression, postnatal depression and parenting stress. BMC Psychiatry, 8, 24.

54. Cole, S. R., & Hernán, M. A. (2002). Fallibility in estimating direct effects. International Journal of Epidemiology, 31, 163–165.

55. Sheikh, M. A., Abelsen, B., & Olsen, J. A. (2016). Differential recall bias, intermediate confounding, and mediation analysis in life course epidemiology: an analytic framework with empirical example. Frontiers in Psychology, 7, 1828. DOI: https://doi.org/10.3389/fpsyg.2016.01828.

56. Loeys, T., Moerkerke, B., Raes, A., Rosseel, Y., & Vansteelandt, S. (2014). Estimation of controlled direct effects in the presence of exposure-induced confounding and latent variables. Structural Equation Modeling: A Multidisciplinary Journal, 21, 396–407.

57. VanderWeele, T. J., & Chiba, Y. (2014). Sensitivity analysis for direct and indirect effects in the presence of exposure-induced mediator-outcome confounders. Epidemiology, Biostatistics, and Public Health, 11, e9027. DOI: 10.2427/9027.

58. De Stavola, B. L., Daniel, R. M., Ploubidis, G. B., & Micali, N. (2015). Mediation analysis with intermediate confounding: structural equation modeling viewed through the causal inference lens. American Journal of Epidemiology, 181, 64–80.

59. Muthén BO, Asparouhov T. (2013). Item response modeling in Mplus: a multi-dimensional, multi-level, and multi-timepoint example. In W. J. van der Linden and R. K. Hambleton, Handbook of item response theory: models, statistical tools, and applications (pp. 1–29). Boca Raton, FL: Chapman & Hall/CRC Press. Available online at: http://www.statmodel.com/download/IRT1Version2.pdf.

60. Hu, L. T., Bentler, P. M. (1998). Fit indices in covariance structure modelling: Sensitivity to underparameterized model misspecification. Psychological Methods, 3, 424–453.

61. Schumacker, R. E., & Lomax, R. G. (2004). A beginner’s guide to structural equation modelling. Mahwah, NJ: Erlbaum.

62. Muthén, L. K., & Muthén, B. O. (2015). Mplus User’s Guide, 7^th^ ed. Muthén & Muthén:Los Angeles, CA.

63. MacKinnon, D. P., Lockwood, C. M, & Williams J. (2004). Confidence limits for the indirect effect: distribution of the product and resampling methods. Multivariate Behavioural Research, 39, 99–128.

64. Tanquerel, S., & Grau-Grau, M. (2020). Unmasking work-family balance barriers and strategies among working fathers in the workplace. Organization, 27, 680–700.

65. Dermott, E. (2005). Time and labour: Fathers’ perceptions of employment and childcare. The Sociological Review, 53, 89–103.

66. Kane, P., & Garber, J. (2004). The relations among depression in fathers, children’s psychopathology, and father–child conflict: A meta-analysis. Clinical Psychology Review, 24, 339–360.

67. Kane, P., & Garber, J. (2009). Parental depression and child externalizing and internalizing symptoms: Unique effects of fathers’ symptoms and perceived conflict as a mediator. Journal of Child and Family Studies, 18, 465–472.

68. Gutierrez-Galve, L., Stein, A., Hanington, L., Heron, J., & Ramchandani, P. (2015). Paternal depression in the postnatal period and child development: mediators and moderators. Pediatrics, 135(2), e339–e347.

69. Connell, A. M., & Goodman, S. H. (2002). The association between psychopathology in fathers versus mothers and children’s internalizing and externalizing behavior problems: a meta-analysis. Psychological Bulletin, 128, 746–773.

70. Matthey, S., Barnett, B., Ungerer, J., & Waters, B. (2000). Paternal and maternal depressed mood during the transition to parenthood. Journal of Affective Disorders, 60, 75–85.

71. Paulson, J. F., & Bazemore, S. D. (2010). Prenatal and postpartum depression in fathers and its association with maternal depression: a meta-analysis. JAMA Network, 303, 1961–1969.

72. Sweeney, S., & MacBeth, A. (2016). The effects of paternal depression on child and adolescent outcomes: a systematic review. Journal of Affective Disorders, 205, 44–59.

73. Schacht, P. M., Cummings, E. M., & Davies, P. T. (2009). Fathering in family context and child adjustment: A longitudinal analysis. Journal of Family Psychology, 23(6), 790.

74. Kerig, P. K. (2019). Parenting and family systems. In M. H. Bornstein (Eds.), Handbook of parenting. Vol. 3. Being and becoming a parent (3rd ed., pp. 3–35). New York: Routledge. DOI: https://www.taylorfrancis.com/books/e/9780429433214/chapters/10.4324/9780429433214-1

75. Dermott, E., & Miller, T. (2015). More than the sum of its parts? Contemporary fatherhood policy, practice and discourse. Families, Relationships and Societies, 4, 183–195.

76. Miller, T. (2010). “It’s a triangle that’s difficult to square": Men’s intentions and practices around caring, work and first-time fatherhood. Fathering, 8, 362.

77. Bornstein, M. H. Toward a model of culture↔parent↔child transactions. In A. Sameroff (Eds.), The Transactional Model of Development: How Children and Contexts Shape Each Other (pp. 139–161). Washington, DC: American Psychological Association, 2009. DOI: http://dx.doi.org/10.1037/11877-000.

78. Cicchetti, D., Toth, S. L., Luthar, S. S., Burack, J. A., & Weisz, J. R. (1997). Developmental psychopathology: Perspectives on adjustment. Cambridge, UK: Cambridge University Press.

79. Sameroff, A. (2009). The transactional model of development: How children and contexts shape each other. Washington, DV: American Psychological Association.

80. Xerxa, Y., Rescorla, L. A., van der Ende, J., Hillegers, M. H., Verhulst, F. C., & Tiemeier, H. (2020). From Parent to Child to Parent: Associations Between Parent and Offspring Psychopathology. Child Development, 92, 291–307.

81. Boyle, M. H., & Pickles, A. (1997). Maternal depressive symptoms and ratings of emotional disorder symptoms in children and adolescents. Journal of Child Psychology and Psychiatry, 38, 981–992.

82. Ordway, M. R. (2011). Depressed mothers as informants on child behavior: Methodological issues. Research in Nursing & Health, 34, 520–532.

83. De Los Reyes, A., & Kazdin, A. E. (2005). Informant discrepancies in the assessment of childhood psychopathology: a critical review, theoretical framework, and recommendations for further study. Psychological Bulletin, 131, 483–509.

84. Burt, K. B., Van Dulmen, M. H., Carlivati, J., Egeland, B., Alan Sroufe, L., Forman, D. R., Appleyard, K., & Carlson, E. A. (2005). Mediating links between maternal depression and offspring psychopathology: The importance of independent data. Journal of Child Psychology and Psychiatry, 46(5), 490–499.

85. Hammen, C., Brennan, P. A., & Shih, J. H. (2004). Family discord and stress predictors of depression and other disorders in adolescent children of depressed and nondepressed women. Journal of the American Academy of Child & Adolescent Psychiatry, 43, 994–1002.

86. Cummings, M.E., Keller, P. S., & Davies, P. T. (2005). Towards a family process model of maternal and paternal depressive symptoms: Exploring multiple relations with child and family functioning. Journal of Child Psychology and Psychiatry, 46, 479–489.

87. Brandon, A. R., Ceccotti, N., Hynan, L. S., Shivakumar, G., Johnson, N., & Jarrett, R. B. (2012). Proof of concept: Partner-Assisted Interpersonal Psychotherapy for perinatal depression. Archives of women’s Mental Health, 15(6), 469–480.

88. Ingram, J., Johnson, D., O’Mahen, H., Kessler, D., Taylor, F.H., Law, R., Round, J., Ford, J., Hopley, R., Culpin, I., & Evans, J. (2019). Protocol for a feasibility randomised trial of low-intensity interventions for antenatal depression: ADAGIO trial comparing interpersonal counselling with cognitive behavioural therapy. BMJ Open, 9(8), e032649. DOI: http://dx.doi.org/10.1136/bmjopen-2019-032649

89. Balldin, S., Fisher, P. AG., & Wirtberg, I. (2018). Video feedback intervention with children: a systematic review. Research on Social Work Practice, 28(6), 682–695.

90. Lawrence, P. J., Davies, B., & Ramchandani, P. G. (2013). Using video feedback to improve early father–infant interaction: A pilot study. Clinical Child Psychology and Psychiatry, 18(1), 61–71.

91. Olhaberry, M., León, M. J., Sieverson, C., Escobar, M., Iribarren, D., Morales-Reyes, I., Mena, C., & Leyton, F. (2019). Is it possible to improve early childhood development with a video-feedback intervention directed at the mother-father-child triad? Research in Psychotherapy: Psychopathology, Process, and Outcome, 22(2), 324. DOI: 10.4081/ripppo.2019.324

92. Cowan, P. A., & Cowan, C. P. (2006). Developmental psychopathology from family systems and family risk factors perspectives: Implications for family research, practice, and policy. In D. Cicchetti, D.J. Cohen (eds.). Developmental Psychopathology. New York: Wiley.

93. Mchale, J. P. (2007). When infants grow up in multiperson relationship systems. Infant Mental Health Journal, 28(4), 370–392.

94. Bauer, A., Knapp, M., & Parsonage, M. (2016). Lifetime costs of perinatal anxiety and depression. Journal of Affective Disorders, 192, 83–90.

