## Supplementary Material for "Maternal postnatal depression and offspring emotional and behavioural development at age 7 years in a UK-birth cohort: the role of paternal involvement"

*Measures*

*Mediators: child-focused and mother-influenced dimensions of paternal involvement*

Potential parenting items (>150) were extracted from paternal self-reported questionnaires administered on 5 occasions after the birth of the study child (8 weeks and 8 months postnatally, 1 year 9 months, 2 years 9 months, 3 years 11 months). We specifically focused on parenting items collected during the first four years of child’s life to capture possible early mechanisms of familial transmission of depression through paternal parenting and quality of father-child relationship. Extracted items captured various dimensions of paternal involvement, which were double-rated and independently assigned into theoretical dimensions by three researchers (IC, RP and TC) in the first instance. This iterative process was followed by extensive discussions in a larger research group with significant input from experts in early child development and parenting (AS and MB). Conceptual factor underpinnings were drawn from extensive empirical and sociological literature on paternal involvement in early infancy.^1-5^ In line with revised conceptualisation of paternal involvement,^4-6^ individual items fell into two theoretically distinct sources of paternal involvement: (1) *child-focused* paternal involvement capturing behavioural (e.g., direct involvement in caregiving), affective (e.g., quality of father-child relationship, such as warmth/enjoyment and conflict, worries about the child) and cognitive (e.g., parenting confidence and beliefs regarding caregiving) dimensions directed at the child; and (2) *mother-influenced* paternal involvement with the child through the lens of maternal expectations (e.g., maternal ‘gatekeeping’, managing employment and parenthood), mother-father relationship (e.g., paternal beliefs regarding mother-father relationship and its impact on parenting) and indirect material care through support of the mother (e.g., paternal help with household tasks and responsibilities). The mother-influenced dimensions of paternal involvement acknowledge those aspects of paternal involvement that are done for the child but do not entail interactions with the child, as well as the impact of inter-parental relationship on parent-child relationship.^5,7^ It has been argued that men’s engagement and interactions with their children need to be understood through the lens of maternal influences^3-5^, with fathers being more involved when their female partners have supportive attitudes regarding paternal involvement, and less involved when inter-parental relationship is characterised by underlying conflict, marital dissatisfaction and maternal ‘gatekeeping’^8^ (maternal beliefs and practices that discourage or facilitate paternal involvement).^9-11^ Evidence suggests that such influences may be more pronounced for paternal rather maternal involvement and parenting behaviours.^12-14^ Supporting the mother through sharing household tasks and responsibilities is another important dimension of paternal mother-influenced involvement.^6^ It may not directly land itself into an aspect of involvement, however, variations in this behaviour may have important implications for the child.^10,11^ It has been acknowledged that similar to mothers, fathers are also under pressure to balance caregiving with demands of working life and traditional ‘breadwinning’ roles,^15-17^ often accompanied by mothers’ perceptions that fathers are not ‘pulling their fair share’ with this ‘benchmarking’ influencing paternal involvement.^3^

The final factors capturing child-focused paternal involvement (Table 1) encompassed: behavioural (e.g., direct involvement in caregiving), affective (e.g., quality of father-child relationship, such as enjoyment and warmth, conflictual relationship with and worries about the child), and cognitive (e.g., paternal parenting confidence and beliefs regarding caregiving) dimensions. The final factors capturing mother-influenced paternal involvement (Table 2) included: paternal perceptions of maternal beliefs or practices that discourage or facilitate his involvement in childcare (e.g., maternal ‘gatekeeping’), paternal help with household tasks and responsibilities (e.g., preparing meals and cleaning home), paternal beliefs regarding mother-father relationship and its impact on parenting (e.g., partner feels hurt by attention mother gives child) and maternal expectations around employment and parenthood (e.g., mother expects partner to take child after work).

*Potential baseline confounder: child polygenic score for neuroticism*

Analyses were adjusted for child polygenic score (PGS) for neuroticism to account for possible child genetic correlations including evocative associations with parenting.^18^ Genotyped data were available for 8,237 children in the ALSPAC cohort. Previously, 116 independent variants robustly associated with neuroticism have been identified.^19^ Of these original variants, 109 were available in the ALSPAC participants with the genetic data. Neuroticism PGS scores were calculated as the sum of the number of copies of each effect allele carried by an individual (ranging from 0 to 2 for each SNP), multiplied by the effect estimate identified in the original GWAS. These weighted neuroticism PGS sum scores were then standardised prior to being included into the analyses.

*Potential intermediate confounder: paternal postnatal depression*

We accounted for possible baseline antenatal confounders of the exposure-outcome, exposure-mediator and mediator-outcome associations by including them in the regression models to estimate each of these pathways.^20^ However, evidence suggests that maternal depression in pregnancy and during the postnatal period is associated with increased risk of paternal PND,^21-22^ which in turn influences paternal parenting,^23^ and offspring emotional and behavioural development,^24^ acting as a potential exposure induced intermediate confounder of the mediator-outcome association.^25,26^ Failure to account for intermediate confounders may result in biased inferences regarding direct and indirect (mediated) effects;^27-29^ thus, we accounted for paternal PND assessed using the Edinburgh Postnatal Depression Scale (EPDS)^30^ administered to fathers at 8 months after the birth of the study child. The EPDS is a 10-item self-reported depression questionnaire, validated and used extensively to screen for depression during the perinatal period in women and men.^31,32^ As with maternal PND, individual items were summed-up to derive a continuous measure (score range 0-24, with higher scores indicating more severe depressive symptoms) to capture the full variation in depressive symptoms.

**Statistical analysis**

*Latent factor models*

Individual parenting items that were primarily theoretically relevant and had standardised loadings of >0.15 were loaded onto the hypothesised parenting dimensions and modelled using Confirmatory Factor Analyses (CFA), a subset of SEM with a robust Weighted Least Square (WLSMV) estimator in M*plus* recommended to model both categorical and continuous data.^33^ To identify the model, the mean of the latent factors was fixed to ‘0’ and their variance to ‘1’. The majority of individual items loaded strongly onto the factors that they were initially theoretically assigned to. However, some items were moved between the factors if modification indices from the initial model indicated that items would be a better fit on the alternative factor, and this made theoretical sense following discussion by the research group. Additional correlations between similar items that were collected at different time points were added into the models to account for shared variance related to time and the repeated nature of the questions. Derived factors, items and factor loadings for each of the measurement models (child-focused and mother-influenced paternal involvement) are presented in Tables 1-2. The Root Mean Square Error of Approximation (RMSEA; >0.06), Comparative Fit Index and Tucker-Lewis Index (CFI/TLI; >0.95) were used to evaluate the fit of the models.^34^

*Direct and mediated effects*

We examined the extent to which the association between maternal PND (8 weeks) and offspring emotional and behavioural development (7 years) is explained, i.e., mediated, by child-focused (six latent factors; Figure 3a) and mother-influenced (four latent factors; Figure 3b) dimensions of paternal involvement using Structural Equation Modelling (SEM) in M*plus* v.8.3.^35^ Analyses of the mediation models were restricted to those with complete data on exposure, outcome, child PGS, baseline and intermediate confounders (n=3,434).

First, we estimated the unadjusted models composed of exposure, outcome and mediators only. Second, we estimated models adjusted for child PGS and all antenatal baseline confounders (Model 1), and further adjusted for paternal PND (8 months) as a possible intermediate confounder of the mediator-outcome association (Model 2), which is also a mediator of the exposure -> mediator association. Thus, our path-specific effects of interest were: (i) Model 1: the indirect effect composed of maternal PND -> paternal involvement -> offspring emotional and behavioural development pathways, and the direct effect, which is the pathway from maternal PND to offspring emotional and behavioural development; (ii) Model 2: the indirect effect composed of a combination of pathways maternal PND -> paternal PND-> paternal involvement -> offspring emotional and behavioural development and maternal PND -> paternal involvement -> offspring emotional and behavioural development pathways, and the direct effect, which is a combination of maternal PND -> paternal PND -> offspring emotional and behavioural development pathways and maternal PND -> offspring emotional and behavioural development pathway. Figure 3a-b represent pathways that constitute total indirect (bold lines) and direct (dashed lines) pathways.

We used MODEL INDIRECT (Model 1) and MODEL CONSTRAINT (Model 2) commands in M*plus* to estimate direct, total indirect and specific indirect effects with a WLSMV estimator to model continuous and categorical variables. Indirect effects [95% CIs] were calculated using product of coefficients method and bias-corrected (BC) bootstrapping (n=1,000 replications) to account for the non-normal distribution.^36^ Results from path analyses with continuous score (offspring total difficulties score), including indirect effects, are presented as unstandardised regression coefficients (hereafter referred to as *β*).

*Missing data: multiple imputation*

We imputed missing data due to loss to follow-up because ignoring those with missing data may result in bias by assuming that data are Missing Completely at Random (MCAR).^37^ We repeated direct and mediated effects analyses on the imputed datasets to account for the loss to follow-up. We used Multivariable Imputation by Chained Equations (MICE)^38^ to impute missing data in exposure, outcome and confounders (except child neuroticism PGS; n=6,029) using the *ice* command in Stata v.15.1/MP (Stata.Corp., Texas, USA). We did not impute child neuroticism PGS from the phenotype data due to restrictions associated with quality control and participant consent. ALSPAC provides a wealth of rich, prospectively collected data on a range of sociodemographic, mental health and developmental variables, which enabled us to account for missing data in exposure, outcome and confounders, as well as factors that explain missingness, which is valid under the Missing-At-Random assumption (MAR).^39^ However, the decision was taken not to impute mediators (child-focused and mother-influenced dimensions of paternal involvement) as there are no sufficient auxiliary data on fathers’ parenting in the ALSPAC cohort to justify the plausibility of MAR assumption. The imputation model was fully compatible with the main analyses. Using binary, ordinal logistic and linear regression models as appropriate, 50 imputed datasets by 10 cycles of regression switching were generated. Monte-Carlo errors were less than 10% of the standard error and FMI values were no larger than 0.5, suggesting that 50 imputed datasets were sufficient.^39^ Each imputation model contained all variables in the substantive analyses along with over 40 auxiliary variables pertaining to maternal and paternal characteristics (e.g., paternal age) and psychopathology, offspring development and mental health, as well maternal and paternal indices of socioeconomic adversity. MICE imputations were carried out in Stata v.15.1/MP (Stata.Corp., Texas, USA). The imputed datasets were exported into M*plus* v.8.3^35^ to estimate direct and indirect (mediated) pathways using MODEL CONSTRAINT command.

**Results S1**

***Sample characteristics***

Characteristics of the study sample and offspring total difficulties mean score at age 7 years by the exposure status (maternal PND at 8 weeks) are presented in Table S1. In summary, mothers who reported higher levels of education (A-Level/University Degree) were more likely to experience depressive symptoms than mothers with lower levels of educational attainment. However, younger, never married mothers, who reported higher levels of inter-parental conflict and financial difficulties, as well as residing in private and/or council rented accommodation were more likely to report depressive symptoms than those mothers who were older, married, did not report experiencing financial difficulties and resided in owned and/or mortgaged accommodation. There was strong evidence that offspring of depressed mothers had higher total difficulties mean scores than those whose mothers did not report experiencing depression during the postnatal period.

***Paternal involvement factors***

Full details of six factors capturing child-focused and four factors capturing mother-influenced aspects of paternal involvement are provided below.

*Child-focused paternal involvement*

*Factor 1 Paternal parenting confidence:* 11 items relating to paternal feelings of confidence in the parenting role and perceptions of the ability to engage effectively in parenting behaviours^36-37^ (e.g., ‘partner feels confident with child’, ‘partner unsure if doing the right thing’, ‘partner happy with the way he brings up child’) were extracted from paternal self-reported questionnaires administered at 8 weeks, 8 months, 1 year 9 months and 2 years 9 months. Higher factor scores represented higher levels of paternal parenting confidence.

*Factor 2 Paternal conflictual relationship with child:* 19 items relating to conflict, harsh disciplining and irritation with the child (e.g., ‘child gets on partner’s nerves’, ‘partner dislikes mess surrounding child’, ‘smacking is the best way to discipline child’) were extracted from paternal self-reported questionnaires administered at 8 weeks, 8 months, 1 year 9 months and 2 years 9 months. Higher factor scores signified lower levels of conflictual parent-child relationship, irritation with the child and less harshness in paternal disciplining.

*Factor 3 Paternal enjoyment and warmth:* 27 items relating to feelings of enjoyment, affection, love and warmth toward the child (e.g., ‘partner enjoys child’, ‘partner feels very close to child’, ‘child gives great joy’) were extracted from paternal self-reported questionnaires administered at 8 weeks, 8 months, 1 year 9 months, 2 years 9 months and 3 years 11 months. Higher factor scores represented more paternal enjoyment, affection and warmth toward the child.

*Factor 4 Paternal involvement in childcare:* 8 items describing frequency of paternal involvement in various aspects of childcare (e.g., ‘frequency partner feeds child per week’, ‘frequency partner puts child to bed per week’, ‘frequency partner bathes child per week’) were extracted from paternal self-reported questionnaires administered at 8 weeks and 8 months. Higher factor scores represented higher frequency of paternal involvement in childcare.

*Factor 5 Paternal worries about child:* 4 items pertaining to paternal worries about the child (e.g., ‘partner worries about child when at work’, ‘partner anxious if someone else looks after child’, ‘partner worries about study child when at work’) were extracted from paternal self-reported questionnaires administered at 1 year 9 months and 2 years 9 months. Higher factor scores represented lower levels of paternal worries about the child.

*Factor 6 Paternal beliefs regarding caregiving:* 6 items relating to paternal parenting principles and practices related to structure, regularity and routine in infant care (e.g., ‘babies should fit into parents’ routine’, ‘babies should be fed when hungry’) and attunement, i.e. responsiveness to infant cues (e.g., ‘babies should be picked up when cry’) were extracted from paternal self-reported questionnaires administered at 8 months and 1 year 9 months. Higher factor scores represented more paternal appreciation of regular routine and higher levels of attunement and responsiveness to child’s cues.

*Mother-focused paternal involvement*

*Factor 1 Paternal help with household tasks:* 12 items related to various aspects of paternal help with household tasks (e.g., ‘partner helped with cleaning home since birth’, ‘partner helped with housework since birth’, ‘partner gave help with preparing meals’) were extracted from paternal self-reported questionnaires administered at 8 weeks and 8 months. Higher factor scores represented higher levels of paternal help with household tasks since the child was born.

*Factor 2 Paternal perception of maternal ‘gatekeeping’:* 7 items relating to paternal perceptions of maternal beliefs and behaviours that encourage or hinder paternal involvement in childcare, i.e., maternal ‘gatekeeping’, (e.g., ‘mother excludes partner from childcare’, ‘partner always getting under mother’s feet, ‘mother dislikes partner being involved with childcare’) were extracted from paternal self-reported questionnaires administered at 8 weeks. Higher factor scores indicated higher levels of paternal perception of being supported by the mother and included in childcare, i.e. less maternal ‘gatekeeping’.

*Factor 3 Paternal beliefs regarding mother-father relationship and parenting:* 3 items relating to paternal beliefs regarding the impact of having a child on father-mother relationship (e.g., ‘parenthood has made partner and mother closer’, ‘mother no longer gives partner attention’) were extracted from paternal self-reported questionnaires administered at 8 weeks. Higher factor scores represented paternal beliefs more concordant with positive changes in the nature of mother-father relationship following the birth of the child.

*Factor 4 Paternal attitudes to employment and parenthood:* 5 items relating to paternal beliefs regarding maternal expectations around employment, childcare and paternal difficulties to manage childcare and employment (e.g., ‘mother expects partner to take child after work’, ‘partner too tired to take child after work’, ‘partner finds it hard to cope with child after work’) were extracted from paternal self-reported questionnaires at 8 weeks and 2 years 9 months. Higher factor scores indicated paternal beliefs concordant with less maternal pressure to look after child after work and less difficulties with managing childcare and employment.

***Associations between dimensions of child-focused and mother-influenced paternal involvement***

In summary, paternal parenting confidence was strongly associated with less conflictual father-child relationship, more paternal enjoyment and involvement in childcare, and less paternal worries about the child (Table S2). Paternal conflictual relationship with the child was associated with less enjoyment and warmth, less involvement in childcare, less appreciation of structure and more worries about the child. Paternal enjoyment and warmth were associated with more involvement in childcare and more paternal appreciation of structure and higher levels of attunement to child’s cues. Paternal worries about the child were associated with less appreciation of structure and less involvement in childcare. Paternal beliefs more concordant with the positive impact of having a child on mother-father relationship were associated with higher levels of paternal help with household tasks and less pressure to manage childcare and employment. Paternal perception of maternal support and inclusion in childcare were associated with higher levels of paternal involvement in household tasks, better ability to manage employment and childcare, and more positive feelings on the changing nature of mother-father relationship and parenting following the birth of the child.

***Associations between maternal PND, child-focused and mother-influenced dimensions of paternal involvement and offspring emotional and behavioural development***

Our pathways of interest were those between maternal PND and offspring emotional and behavioural development at age 7 years through child-focused and mother-influenced dimensions of paternal involvement. Specifically, we examined the pathways from maternal PND to paternal involvement (both direct and through paternal PND), and the pathways from paternal involvement to offspring emotional and behavioural development. Full description of estimates for Model 1 and Model 2 are provided below.

***Exposure (maternal PND) – mediator (paternal involvement) associations***

In Model 1 (adjusted for child PGS and antenatal baseline confounders; Table 1) maternal PND at 8 weeks was strongly associated with less paternal parenting confidence (β=-0.049, 95% CI: -0.061, -0.037, p≤0.001), more father-child conflict (β=-0.053, 95% CI: -0.063, -0.043, p≤0.001), less paternal enjoyment and warmth (β=-0.032, 95% CI: -0.042, -0.022, p≤0.001), more paternal worries about the child (β=-0.027, 95% CI: -0.039, -0.015, p≤0.001), higher levels of perceived maternal ‘gatekeeping’ (β=-0.040, 95% CI: -0.054, -0.026, p≤0.001), more negative feelings regarding the impact the birth of the child have had on mother-father relationship (β=-0.030, 95% CI: -0.042, -0.018, p≤0.001), and paternal feelings of more pressure to look after the child after work and more struggles to manage childcare and employment (β=-0.031, 95% CI: -0.041, -0.021, p≤0.001; Table 3).

In contrast, in Model 2 (further adjusted for paternal PND at 8 months as an intermediate confounder; Table 4), maternal PND was strongly associated with more paternal enjoyment and warmth (β=0.055, 95% CI: 0.031, 0.078, p≤0.001) and direct involvement in childcare (β=0.029, 95% CI: 0.017, 0.041, p≤0.001), more paternal parenting confidence (β=0.029, 95% CI: 0.004, 0.054, p=0.027), less father-child conflict (β=0.034, 95% CI: 0.007, 0.061, p=0.016), more paternal help with household tasks (β=-0.009, 95% CI: -0.001, 0.019, p=0.070), and more paternal struggle to manage childcare and employment (β=-0.010, 95% CI: -0.019, -0.001, p=0.047), although 95% CIs were wide, that was not explained through paternal PND.

Maternal PND also had an indirect effect on paternal engagement via paternal PND (Table 4). Specifically, maternal PND was strongly associated with paternal PND (β=0.139, 95% CI: 0.115, 0.162, p≤0.001), which, in turn, was strongly associated with less paternal parenting confidence (β=-0.812, 95% CI: -0.935, -0.689, p≤0.001), less enjoyment and warmth (β=-0.784, 95% CI: -0.884, -0.684, p≤0.001), less involvement in childcare (β=-0.163, 95% CI: -0.200, -0.128, p≤0.001) and appreciation of regular routine (β=-0.102, 95% CI: -0.141, -0.063, p≤0.001), more father-child conflict (β=-0.975, 95% CI: -1.124, -0.826, p≤0.001) and more paternal worries about the child (β=-0.117, 95% CI: -0.150, -0.084, p≤0.001), less paternal help with household tasks (β=-0.061, 95% CI: -0.083, -0.039, p≤0.001), higher levels of perceived maternal ‘gatekeeping’ (β=-0.366, 95% CI: -0.427, -0.305, p≤0.001), more negative feelings regarding the impact the birth of the child have had on mother-father relationship (β=-0.416, 95% CI: -0.494, -0.338, p≤0.001), and paternal feelings of more pressure to look after the child after work and more struggles to manage childcare and employment (β=-0.169, 95% CI: -0.194, -0.143, p≤0.001; Table 4).

***Mediator (paternal involvement) – outcome (offspring emotional and behavioural development) associations***

In Model 1, lower levels of paternal parenting confidence (β=-0.597, 95% CI: -0.789, -0.404, p≤0.001), warmth and enjoyment (β=-0.379, 95% CI: -0.551, -0.206, p≤0.001), higher levels of father-child conflict (β=-0.779, 95% CI: -0.961, -0.597, p≤0.001) and worries about the child (β=-0.277, 95% CI: -0.479, -0.075, p=0.007) were associated with higher risk of offspring emotional and behavioural difficulties at age 7 years (Table 3). Out of all mother-influenced dimensions of paternal involvement, only paternal feelings of more pressure to look after the child after work and more struggles to manage childcare and employment were associated with increased risk of offspring emotional and behavioural difficulties (β=-0.224, 95% CI: -0.391, -0.057, p=0.008). In Model 2, however, the only child-focused dimension of paternal involvement, higher levels of father-child conflict, were strongly associated with higher risk of offspring emotional and behavioural difficulties at age 7 years (β=-0.546, 95% CI: -0.998, -0.093, p=0.018; Table 4), with no evidence for associations between any of the mother-influenced dimensions of paternal involvement and offspring emotional and behavioural development.

***Direct and mediated effects***

In Model 1 (adjusted for child PGS and antenatal baseline confounders; Table 5), there was evidence of total indirect effect from early maternal PND to offspring development at age 7 years through the combination of all parenting factors capturing child-focused (β=0.090, 95%CIs: 0.063, 0.117, p≤0.001; Table 3), but not mother-influenced dimensions of paternal involvement (β=0.010, 95%CIs: -0.008, 0.028, p=0.291). There was strong evidence of total and direct effects from maternal PND to offspring emotional and behavioural development at age 7 years in models capturing both child-focused (total effect: β=0.210, 95%CIs: 0.171, 0.250, p≤0.001; direct effect: β=0.120, 95%CIs: 0.077, 0.163, p≤0.001) and mother-influenced (total effect: β=0.210, 95%CIs: 0.171, 0.250, p≤0.001; direct effect: β=0.200, 95%CIs: 0.157, 0.243, p≤0.001) paternal involvement. Amongst child-focused dimensions of paternal involvement, there was evidence for specific indirect effects through paternal parenting confidence (β=0.030, 95%CIs: 0.018, 0.042, p≤0.001), paternal conflictual relationship with the child (β=0.041, 95%CIs: 0.029, 0.053, p≤0.001), paternal enjoyment and warmth (β=0.012, 95%CIs: 0.004, 0.020, p≤0.001), and paternal worries about the child (β=0.007, 95%CIs: 0.001, 0.013, p=0.028), but not paternal involvement in childcare and parenting principles and practices. There was only some evidence for specific indirect effect through paternal attitudes to employment and parenthood (β=0.007, 95%CIs: 0.001, 0.013, p=0.032), although 95% CIs were wide, amongst mother-influenced dimensions of paternal involvement.

In contrast, in Model 2 (further adjusted for paternal PND at 8 months as an intermediate confounder; Table 5), there was no evidence of total indirect effect from early maternal PND to offspring development at 7 years through the combination of all factors capturing child-focused (β=0.082, 95%CIs: -0.028, 0.192, p=0.144) and mother-influenced (β=-0.023, 95%CIs: -0.066, 0.020, p=0.282) dimensions of paternal involvement. Similarly to Model 1, there was strong evidence of total and direct effects from maternal PND to offspring emotional and behavioural development at age 7 years in models capturing both child-focused (total effect: β=0.267, 95%CIs: 0.077, 0.457, p≤0.001; direct effect: β=0.185, 95%CIs: 0.110, 0.273, p≤0.001) and mother-influenced (total effect: β=0.163, 95%CIs: 0.092, 0.234, p≤0.001; direct effect: β=0.186, 95%CIs: 0.139, 0.233, p≤0.001) paternal involvement. However, unlike Model 1, there was only some evidence for specific indirect effect through paternal conflictual relationship with the child (β=0.055, 95%CIs: 0.001, 0.110, p=0.046), although the 95% CIs were wide, but not other child-focused dimensions of paternal involvement. There was no evidence of specificity through any of the mother-influenced dimensions of paternal involvement.

***Sensitivity analyses: missing data***

We repeated direct and mediated effects analyses on imputed datasets to examine the impact of response attrition on our findings. Characteristics of the sample by the completeness of the data are presented in Table S3. Participants comprising our analytical sample were from a higher socioeconomic background as indexed by lower proportion of those reporting financial difficulties and higher proportion of those reporting residing owned/mortgaged accommodation and married marital status compared to the original ALSPAC sample. The results from the analyses using the imputed data supported our findings resulting in the similar pattern of results and over-arching conclusions as our main findings in the complete case analyses (Table S4) with one notable difference. In Model 2, there was stronger evidence for the specific indirect effect through paternal conflictual relationship with the child (*B*=0.042, 95%CIs: 0.005, 0.079, p=0.028), with some emerging evidence for a specific indirect effect through paternal worries about the child although the 95%CIs were wide (*B*=0.006, 95%CI 0.001, 0.012, p=0.037) compared to complete case analyses (Table 5).

**References**

1. Dermott, E. (2014). *Intimate fatherhood: A sociological analysis*. Routledge.
2. Lamb, M. E., & Lewis, C. (2004). The development and significance of father–child relationships in two-parent families. In M. E. Lamb (Eds.), The role of the father in child development (4th ed., pp. 272 – 306). Hoboken, NJ: Wile
3. Marsiglio, W., Day, R. D., & Lamb, M. E. (2000). Exploring fatherhood diversity: Implications for conceptualizing father involvement. *Marriage & Family Review*, *29*, 269-293.
4. Palkovitz, R. (2019). Expanding our focus from father involvement to father–child relationship quality. *Journal of Family Theory & Review*, *11*(4), 576-591.
5. Pleck, J. H. (2012). Integrating father involvement in parenting research. *Parenting*, *12*, 243-253.
6. Schoppe-Sullivan, S. J., McBride, B. A., & Ho, M. H. R. (2004). Unidimensional versus multidimensional perspectives on father involvement. *Fathering: A Journal of Theory, Research & Practice about Men as Fathers*, *2*, 147-164.
7. Cabrera, N. J., Fitzgerald, H. E., Bradley, R. H., & Roggman, L. (2014). The ecology of father‐child relationships: An expanded model. *Journal of Family Theory & Review*, *6*(4), 336-354.
8. Miller, T. (2018). Paternal and maternal gatekeeping? Choreographing care. *Sociologica*, *12*, 25-35.
9. Belsky, J., Youngblade, L., Rovine, M., & Volling, B. (1991). Patterns of marital change and parent-child interaction. *Journal of Marriage and the Family*, 487-498.
10. Feldman, R. (2000). Parents' convergence on sharing and marital satisfaction, father involvement, and parent–child relationship at the transition to parenthood. *Infant Mental Health Journal: Official Publication of The World Association for Infant Mental Health*, *21*(3), 176-191.
11. Pleck, J. H. (2010). Paternal involvement: Revised conceptualisation and theoretical linkages with child outcomes. In M. Lamb (Eds.), *The role of the father in child development (pp. 58-93).* Hoboken, NJ: Wiley.
12. Beitel, A. H., & Parke, R. D. (1998). Paternal involvement in infancy: The role of maternal and paternal attitudes. *Journal of Family Psychology*, *12*(2), 268.
13. Belsky, J., Gilstrap, B., & Rovine, M. (1984). The Pennsylvania Infant and Family Development Project, I: Stability and change in mother-infant and father-infant interaction in a family setting at one, three, and nine months. *Child development*, 692-705.
14. Schacht, P. M., Cummings, E. M., & Davies, P. T. (2009). Fathering in family context and child adjustment: A longitudinal analysis. *Journal of family psychology*, *23*(6), 790.
15. Cabrera, N. J., & Tamis-LeMonda, C. S. (Eds.). (2013). *Handbook of father involvement: Multidisciplinary perspectives*. Routledge.
16. Marsiglio, W., Amato, P., Day, R. D., & Lamb, M. E. (2000). Scholarship on fatherhood in the 1990s and beyond. *Journal of marriage and family*, *62*(4), 1173-1191.
17. Miller, T. (2010). *Making sense of fatherhood: Gender, caring and work*. Cambridge University Press.
18. Stein, A., Pearson, R.M., Goodman, S.H., Rapa, E., Rahman, A., McCallum, M., Howard, L.M., & Pariante, C.M. (2014). Effects of perinatal mental disorders on the fetus and child. *The Lancet*, 384, 1800-1819.
19. Luciano M, Hagenaars SP, Davies G, et al. Association analysis in over 329,000 individuals identifies 116 independent variants influencing neuroticism. *Nature Genetics.* 2018;*50*(1):6-11.
20. Northstone, K., Lewcock, M., Groom, A., Boyd, A., Macleod, J., Timpson, N.J., & Wells, N. (2019). The Avon Longitudinal Study of Parents and Children (ALSPAC): an update on the enrolled sample of index children in 2019. *Wellcome Open Research*, 4:51. DOI:
21. Goodman, J. H. (2004). Paternal postpartum depression, its relationship to maternal postpartum depression, and implications for family health. *Journal of Advanced Nursing*, *45*, 26-35.
22. Fredriksen, E., Von Soest, T., Smith, L., & Moe, V. (2019). Depressive symptom contagion in the transition to parenthood: Interparental processes and the role of partner-related attachment. *Journal of Abnormal Psychology*, *128*, 397-403.
23. Wilson, S., & Durbin, C. E. (2010). Effects of paternal depression on fathers' parenting behaviors: A meta-analytic review. *Clinical Psychology Review*, *30*, 167-180.
24. Ramchandani, P., Stein, A., Evans, J., O'Connor, T. G., & ALSPAC Study Team. (2005). Paternal depression in the postnatal period and child development: a prospective population study. *The Lancet*, 365, 2201-2205.
25. Cole, S. R., & Hernán, M. A. (2002). Fallibility in estimating direct effects. *International Journal of Epidemiology*, *31*, 163-165.
26. Sheikh, M. A., Abelsen, B., & Olsen, J. A. (2016). Differential recall bias, intermediate confounding, and mediation analysis in life course epidemiology: an analytic framework with empirical example. *Frontiers in Psychology*, *7*, 1828. DOI: <https://doi.org/10.3389/fpsyg.2016.01828>.
27. De Stavola, B. L., Daniel, R. M., Ploubidis, G. B., & Micali, N. (2015). Mediation analysis with intermediate confounding: structural equation modeling viewed through the causal inference lens. *American Journal of Epidemiology*, *181*, 64-80.
28. Loeys, T., Moerkerke, B., Raes, A., Rosseel, Y., & Vansteelandt, S. (2014). Estimation of controlled direct effects in the presence of exposure-induced confounding and latent variables. *Structural Equation Modeling: A Multidisciplinary Journal*, *21*, 396-407.
29. VanderWeele, T. J., & Chiba, Y. (2014). Sensitivity analysis for direct and indirect effects in the presence of exposure-induced mediator-outcome confounders. *Epidemiology, Biostatistics, and Public Health*, *11,* e9027. DOI:  [10.2427/9027](https://dx.doi.org/10.2427%2F9027).
30. Cox, J. L., Holden, J. M., & Sagovsky, R. (1987). Detection of postnatal depression: development of the 10-item Edinburgh Postnatal Depression Scale. *British Journal of Psychiatry*, *150*, 782-786.
31. Massoudi, P., Hwang, C. P., & Wickberg, B. (2013). How well does the Edinburgh Postnatal Depression Scale identify depression and anxiety in fathers? A validation study in a population based Swedish sample. *Journal of affective disorders*, *149*(1-3), 67-74.
32. Matthey, S., Barnett, B., Kavanagh, D. J., & Howie, P. (2001). Validation of the Edinburgh Postnatal Depression Scale for men, and comparison of item endorsement with their partners. *Journal of affective disorders*, *64*(2-3), 175-184.
33. Muthén BO, Asparouhov T. (2013). Item response modeling in Mplus: a multi-dimensional, multi-level, and multi-timepoint example. In W. J. van der Linden and R. K. Hambleton, *Handbook of item response theory: models, statistical tools, and applications* (pp. 1-29).

Boca Raton, FL: Chapman & Hall/CRC Press. Available online at: <http://www.statmodel.com/download/IRT1Version2.pdf>.

34. Hu, L. T., Bentler, P. M. (1998). Fit indices in covariance structure modelling: Sensitivity to underparameterized model misspecification. *Psychological Methods*, 3, 424-453.
35. Muthén, L. K., &Muthén, B. O. (2015). *Mplus User’s Guide*, 7^th^ ed. Muthén & Muthén:Los Angeles, CA.
36. MacKinnon, D. P., Lockwood, C. M, & Williams J. (2004). Confidence limits for the indirect effect: distribution of the product and resampling methods. *Multivariate Behavioural Research*, 39, 99-128.
37. Sterne JAC, White IR, Carlin JB, et al. Multiple imputation for missing data in epidemiological and clinical research: potential and pitfalls. *Brit Med J*. 2009;338:b2393-b2393.
38. Royston P, White IR. Multiple imputation by chained equations (MICE): implementation in Stata. Journal of Statistical Software. 2011;45:1-20.
39. White IR, Royston P, Wood AM. Multiple imputation using chained equations: issues and guidance for practice. *Stat Med*. 2011;30(4):377-99.

**Table S1.** Characteristics of the sample and offspring total difficulties mean score at age 7 years by the exposure status (maternal postnatal depression (PND) at 8 weeks dichotomised at a cut-off ≥13)

| Exposure status:  N  (%) | Maternal postnatal depression (8 weeks) | |
| --- | --- | --- |
|  | No  10,383  (89.9) | Yes  1,170  (10.1) |
|  | n (%) | n (%) |
| *Maternal educational attainment* |  |  |
| A-Level/Degree | 5,719 (89.8) | 653 (10.2) |
| Minimal education/None/O-Level | 3,718 (91.4) | 349 (8.6) |
| Chi^2^, p-value |  | 7.95, 0.005 |
| *Financial difficulties* |  |  |
| No financial difficulties | 8,183 (91.8) | 733 (8.2) |
| Financial difficulties | 1,934 (82.9) | 399 (17.1) |
| Chi^2^, p-value |  | 161. 16, <0.001 |
| *Child gender* |  |  |
| Male | 5,353 (89.8) | 610 (10.2) |
| Female | 5,030 (90.0) | 560 (10.0) |
| Chi^2^, p-value |  | 0.14, 0.706 |
| *Home ownership* |  |  |
| Owned/mortgaged | 7,546 (91.9) | 662 (8.1) |
| Private/council rented | 1,692 (84.5) | 310 (15.5) |
| Chi^2^, p-value |  | 102.85, <0.001 |
| *Marital status* |  |  |
| Never married | 2,181 (85.9) | 359 (14.1) |
| Married | 7,886 (91.2) | 760 (8.8) |
| Chi^2^, p-value |  | 62.27, <0.001 |
| *Maternal age, mean (SD)* | 27.6 (4.8) | 27.2 (5.2) |
| ANOVA, p-value |  | 2.88, 0.004 |
| *Offspring total difficulties score at 7 years* | 7.3 (4.7) | 9.8 (5.6) |
| ANOVA, p-value |  | -13.58, <0.001 |
| *Parental antenatal conflict* | 9.2 (1.9) | 10.1 (1.7) |
| ANOVA, p-value |  | 16.14, <0.001 |

*Note*: p-values based on Pearson’s Chi-square (Chi^2^) test of association between maternal postnatal depression and categorical variables, and ANOVA for differences in means for continuous variables; sample sizes vary due to the differences in data availability on socioeconomic, child, parental and familial characteristics.

**Table S2.** Associations between child-focused and mother-influenced dimensions of paternal involvement

| Paternal involvement factors | | Point estimate (β)^a^ | S.E. | p-value |
| --- | --- | --- | --- | --- |
| **Six specific factors capturing child-focused dimensions of paternal involvement** | | | | |
| *Paternal parenting confidence* | Paternal conflictual relationship with child | 0.682 | 0.014 | ≤0.001 |
|  | Paternal enjoyment and warmth | 0.687 | 0.013 | ≤0.001 |
|  | Paternal beliefs regarding caregiving | -0.001 | 0.025 | 0.836 |
|  | Paternal involvement in childcare | 0.241 | 0.017 | ≤0.001 |
|  | Paternal worries about child | 0.294 | 0.020 | ≤0.001 |
| *Paternal conflictual relationship with child* | Paternal enjoyment and warmth | 0.695 | 0.011 | ≤0.001 |
|  | Paternal appreciation of routine | 0.057 | 0.023 | 0.017 |
|  | Paternal involvement in childcare | 0.077 | 0.018 | ≤0.001 |
|  | Paternal worries about child | 0.277 | 0.018 | ≤0.001 |
| *Paternal enjoyment and warmth* | Paternal appreciation of regular routine | 0.223 | 0.022 | ≤0.001 |
|  | Paternal involvement in childcare | 0.260 | 0.016 | ≤0.001 |
|  | Paternal worries about child | 0.002 | 0.019 | 0.904 |
| *Paternal beliefs regarding caregiving* | Paternal involvement in childcare | 0.066 | 0.024 | 0.006 |
|  | Paternal worries about child | -0.116 | 0.026 | ≤0.001 |
| *Paternal involvement in childcare* | Paternal worries about child | -0.062 | 0.020 | 0.002 |
| **Four specific factors capturing mother-influenced dimensions of paternal involvement** | | | | |
| *Paternal perceptions of maternal ‘gatekeeping’* | Paternal beliefs regarding mother-father relationship and parenting | 0.740 | 0.020 | ≤0.001 |
|  | Paternal help with household tasks | 0.209 | 0.018 | ≤0.001 |
|  | Paternal beliefs regarding employment and parenthood | 0.283 | 0.017 | ≤0.001 |
| *Paternal beliefs regarding mother-father relationship and parenting* | Paternal help with household tasks | 0.165 | 0.019 | ≤0.001 |
|  | Paternal attitudes to employment and parenthood | 0.302 | 0.017 | ≤0.001 |
| *Paternal help with household tasks* | Paternal beliefs regarding employment and parenthood | -0.002 | 0.014 | 0.898 |

Note: ^a^ Effect size are standardised (the variance of the latent factors was fixed to ‘1’) regression coefficients (*β)*

**Table S3.** Distribution of sociodemographic characteristics in the original Avon Longitudinal Study of Parents and Children (ALSPAC) Cohort and the study complete and imputed samples

| Sample demographic characteristics assessed during pregnancy ^1^ | Core ALSPAC sample ^a^  (n=14,901) | Complete sample ^b^  (n=3,434) | Imputed sample ^c^  (n=6,029) |
| --- | --- | --- | --- |
|  | (%) | (%) | (%) |
| *Maternal educational attainment* |  |  |  |
| A-Level/Degree | 62.4 | 52.0 | 56.7 |
| Minimal education/none/O-Level | 37.6 | 48.0 | 43.3 |
| *Presence of financial difficulties* |  |  |  |
| Financial difficulties | 78.8 | 85.4 | 82.7 |
| No financial difficulties | 21.2 | 14.6 | 17.3 |
| *Homeownership status* |  |  |  |
| Owned/mortgaged | 79.0 | 89.7 | 84.3 |
| Private/council rented | 21.0 | 10.3 | 15.7 |
| *Marital status* |  |  |  |
| Married | 75.0 | 86.9 | 17.3 |
| Never married | 25.0 | 13.1 | 82.7 |
|  | Mean (SD) | Mean (SD) | Mean (SD) |
| *Parental conflict* | 10.01 (1.77) | 10.27 (1.66) | 10.13 (1.70) |
| *Maternal age* | 27.23 (4.96) | 28.73 (4.28) | 28.09 (4.59) |
| *Paternal PND* | 3.35 (3.70) | 3.18 (3.50) | 3.41 (3.67) |

*Note*: ^1^ Additional missing data on demographics: maternal educational attainment missing for 3,237/21.7%; maternal age missing for 931/6.0%; financial difficulties missing for 2,292/14.6%; homeownership status missing for 4,699/30.0%; marital status missing for 2,092/13.4%; parental conflict missing for 3,562/22.8%; paternal PND missing for 8,572/54.8%.

^a^ Core ALSPAC sample: no exposure or outcome data; ^b^ Complete sample: exposure, outcomes and confounders data available; ^c^ Imputed sample: imputed missing data on exposure, outcomes and confounders (except child PGS).

PGS: Polygenic Score for Neuroticism; Paternal PND: Paternal Postnatal Depression.

**Table S4.** Estimates of direct and mediated effects in the unadjusted model and models adjusted for child PRS, gender, antenatal baseline confounders, and paternal PND as an intermediate confounder in imputed sample (n=6,029)

|  | Model estimates (n=6,029) | | | | | |
| --- | --- | --- | --- | --- | --- | --- |
| Effect Size ^1^ | Unadjusted model | | Model 1^a^ | | Model 2^b^ | |
|  | *Β* [95% CI] | P-value | *Β* [95% CI] | P-value | *Β* [95% CI] | P-value |
| **Child-focused dimensions of paternal involvement** | | | | | | |
| *1. Total indirect effect^*^* | 0.098 [0.074, 0.121] | ≤0.001 | 0.083 [0.061, 0.104] | ≤0.001 | 0.054 [-0.030, 0.138] | 0.209 |
| *2. Direct effect^**^* | 0.142 [0.110, 0.175] | ≤0.001 | 0.118 [0.085, 0.151] | ≤0.001 | 0.166 [0.088, 0.244] | ≤0.001 |
| *3. Total effect ^***^* | 0.240 [0.212, 0.267] | ≤0.001 | 0.201 [0.173, 0.228] | ≤0.001 | 0.220 [0.061, 0.379] | 0.006 |
| *4. Specific indirect effects* |  |  |  |  |  |  |
| *Paternal parenting confidence* | 0.027 [0.019, 0.035] | ≤0.001 | 0.025 [0.017, 0.033] | ≤0.001 | 0.012 [-0.023, 0.047] | 0.491 |
| *Paternal conflictual relationship with child* | 0.042 [0.030, 0.054] | ≤0.001 | 0.038 [0.028, 0.048] | ≤0.001 | 0.042 [0.005, 0.079] | 0.028 |
| *Paternal enjoyment and warmth* | 0.014 [0.010, 0.020] | ≤0.001 | 0.012 [0.006, 0.018] | 0.001 | -0.005 [-0.025, 0.015] | 0.625 |
| *Paternal involvement in childcare* | 0.001 [-0.001, 0.003] | 0.488 | 0.001 [-0.001, 0.003] | 0.352 | 0.001 [-0.001, 0.003] | 0.602 |
| *Paternal beliefs regarding caregiving* | 0.001 [-0.001, 0.003] | 0.634 | 0.001 [-0.001, 0.003] | 0.832 | -0.001 [-0.005, 0.003] | 0.520 |
| *Paternal worries about child* | 0.014 [0.006, 0.022] | ≤0.001 | 0.008 [0.002, 0.014] | 0.005 | 0.006 [0.001, 0.012] | 0.037 |
| **Mother-influenced dimensions of paternal involvement** | | | | | | |
| *1. Total indirect effect* | 0.021 [0.005, 0.037] | 0.006 | 0.002 [-0.010, 0.014] | 0.716 | -0.014 [-0.034, 0.006] | 0.193 |
| *2. Direct effect^**^* | 0.222 [0.193, 0.251] | ≤0.001 | 0.197 [0.166, 0.228] | ≤0.001 | 0.191 [0.162, 0.220] | ≤0.001 |
| *3. Total effect ^***^* | 0.243 [0.215, 0.270] | ≤0.001 | 0.199 [0.171, 0.226] | ≤0.001 | 0.177 [0.086, 0.148] | ≤0.001 |
| *4. Specific indirect effects* |  |  |  |  |  |  |
| *Paternal help with household tasks* | 0.001 [-0.001, 0.003] | 0.535 | 0.001 [-0.001, 0.003] | 0.784 | -0.005 [-0.017, 0.007] | 0.397 |
| *Paternal perception of maternal ‘gatekeeping’* | 0.015 [0.003, 0.027] | 0.007 | 0.002 [-0.010, 0.014] | 0.649 | -0.008 [-0.020, 0.004] | 0.191 |
| *Paternal beliefs regarding mother-father relationship and parenting* | 0.001 [-0.007, 0.009] | 0.687 | 0.001 [-0.005, 0.007] | 0.941 | 0.001 [-0.001, 0.003] | 0.761 |
| *Paternal beliefs regarding employment and parenthood* | 0.005 [0.001, 0.010] | 0.006 | 0.001 [-0.001, 0.003] | 0.726 | 0.001 [-0.001, 0.003] | 0.994 |

*Note****:***^1^ Effect size are unadjusted and adjusted regression coefficients (*B* unstandardised); unadjusted model (exposure, outcome and mediators only); Model 1: adjusted for child PGS and antenatal baseline confounders (child gender, financial problems, homeownership status, maternal age and education, parental conflict, marital status); Model 2: further adjusted for paternal PND at 8 months as an intermediate confounder.

**Model 1:**^*^ Total indirect effect = maternal PND->aspects of paternal engagement->offspring emotional and behavioural development; ^**^Direct effect = maternal PND-> offspring emotional and behavioural development; ^***^Total effect = total indirect effect + direct effect.

**Model 2:**^*^ Total indirect effect = maternal PND->aspects of paternal engagement->offspring emotional and behavioural development + maternal PND->paternal PND->aspects of paternal engagement -> offspring emotional and behavioural development; ^**^Direct effect = maternal PND-> offspring emotional and behavioural development + maternal PND->paternal PND->offspring emotional and behavioural development; ^***^Total effect = total indirect effect + direct effect.

Maternal PND: maternal postnatal depression; PGS: Polygenic Score for Neuroticism; Paternal PND: paternal postnatal depression.
